# *LRRK2* coding variants and the risk of Parkinson’s disease

**DOI:** 10.1101/2021.04.22.21255928

**Authors:** Julie Lake, Xylena Reed, Rebekah G. Langston, Mike A. Nalls, Ziv Gan-Or, Mark R. Cookson, Andrew B. Singleton, Cornelis Blauwendraat, Hampton L. Leonard, on behalf of the International Parkinson’s Disease Genomics Consortium (IPDGC)

**Author notes:** Denotes equal contribution (Cornelis Blauwendraat, Hampton L. Leonard). **Correspondence to:** Hampton Leonard, Laboratory of Neurogenetics, NIA, NIH, Building 35, 35 Convent Drive, Bethesda, MD 20892, USA. **Funding sources for study:** This work was supported in part by the Intramural Research Programs of the National Institute of Neurological Disorders and Stroke (NINDS), the National Institute on Aging (NIA), and the National Institute of Environmental Health Sciences both part of the National Institutes of Health, Department of Health and Human Services; project numbers 1ZIA-NS003154, Z01-AG000949-02 and Z01-ES101986.

## Abstract

**Background:** The leucine-rich repeat kinase 2 (*LRRK2*) gene harbors both rare highly damaging missense variants (e.g. p.G2019S) and common non-coding variants (e.g. rs76904798) with lower effect sizes that are associated with Parkinson’s disease risk.

**Objectives:** This study aimed to investigate in a large meta-analysis whether the *LRRK2* GWAS signal represented by rs76904798 is independently associated with Parkinson’s disease risk from *LRRK2* coding variation, and whether complex linkage disequilibrium structures with p.G2019S and the 5’ non-coding haplotype account for the association of *LRRK2* coding variants.

**Methods:** We performed a meta-analysis using imputed genotypes from 17,838 cases, 13,404 proxy-cases and 173,639 healthy controls of European ancestry. We excluded carriers of p.G2019S and/or rs76904798 to clarify the role of *LRRK2* coding variation in mediating disease risk, and excluded carriers of relatively rare *LRRK2* coding variants to assess the independence of rs76904798. We also investigated the co-inheritance of *LRRK2* coding variants with p.G2019S, rs76904798 and p.N2081D.

**Results:** *LRRK2* rs76904798 remained significantly associated with Parkinson’s disease after excluding carriers of relatively rare *LRRK2* coding variants. *LRRK2* p.R1514Q and p.N2081D were frequently co-inherited with rs76904798 and the allele distribution of p.S1647T significantly changed among cases after removing rs76904798 carriers.

**Conclusions:** These data suggest that the *LRRK2* coding variants previously linked to Parkinson’s disease (p.N551K, p.R1398H, p.M1646T and p.N2081D) do not drive the 5’ non-coding GWAS signal. These data, however, do not preclude the independent association of the haplotype p.N551K-p.R1398H and p.M1646T with altered disease risk.

## Introduction

The leucine-rich repeat kinase 2 (*LRRK2*) gene has been a focus of Parkinson’s disease (PD) research since the discovery that pathogenic mutations in the gene are linked to autosomal-dominant PD ^1,2^. The most common pathogenic mutation, p.G2019S, has a relatively high frequency in Ashkenazi Jews and North African Arabs and is also found in ∼1% of Europeans with PD ^3^. Although the underlying mechanism of toxicity is not fully elucidated, the p.G2019S mutation has been shown to increase LRRK2 kinase activity, which is thought to cause a toxic gain of function ^4^. Hyperactive mutations in *LRRK2* have been shown to impact lysosomal and endocytic regulation by increasing *LRRK2* recruitment to lysosomes ^5^. Some neuropathological changes and clinical features caused by LRRK2-PD are similar to those of idiopathic PD, suggesting that there could be common mechanisms of pathogenesis and potentially common therapies ^6^.

Several studies have nominated *LRRK2* missense variants as either causal for or associated with an increased risk of PD in individuals of Asian (e.g. p.A419V, p.R1628P and p.G2385R), Arab-Berber (e.g. p.Y2189C), and European ancestry (e.g. p.M1646T, p.N2081D and p.R1441C/G/H) ^7–14^.A common protective haplotype (p.N551K-p.R1398H-p.K1423K) has also been observed in individuals of both Asian and European ancestry ^7^. In addition, common non-coding variation (rs76904798, OR: 1.15, 95% CI: 1.13-1.18, P=1.52e-28) upstream of *LRRK2* is associated with increased PD risk, an effect that is independent of the p.G2019S mutation ^15^.

Although the exact causal variant at the 5’ GWAS signal of *LRRK2* is unknown ^16,17^, it has been shown in a relatively small cohort of 1381 PD cases and 1328 controls that the 5’ signal had a low degree of correlation with known coding susceptibility variants, including p.M1646T and protective haplotype p.N551K-R1398H-K1423K, indicating the independence of this GWAS signal ^18^. The independence of this signal from *LRRK2* coding variation suggests that changes in the expression or splicing of *LRRK2* could mediate PD risk. There is evidence that the GWAS-nominated non-coding variation tagged by rs76904798 could affect *LRRK2* expression. The allele rs76904798-T has been associated with increased *LRRK2* expression in monocytes ^19^, monocyte-derived microglia-like cells ^20^ and in both human brain microglia and IPSC-derived models ^21^. The same allele has also been associated with faster development of motor symptoms ^22^. To further investigate the pattern of association at the *LRRK2* locus and determine in a significantly larger dataset whether rs76904798 is independently associated with PD from *LRRK2* coding variation, we performed a conditional association analysis using individual-level genotype data from 17,838 PD cases, 13,404 proxy-cases (i.e. individuals with a first degree relative who has PD but do not have PD themself) and 173,639 healthy controls of European ancestry.

## Methods

### Genotyping data

#### International Parkinson Disease Genomics Consortium genotyping data

Genotype data were obtained as previously described ^15^. Only datasets with participants that had high-quality imputation scores (R^2^ □ > □ 0.8) for p.G2019S and the 5’ non-coding variant rs76904798 were included. Quality control parameters can be found in the Supplemental Methods. In total, 13 datasets were included from IPDGC with individual level genotypes from 16,309 PD patients and 17,705 healthy controls with European ancestry (Supplementary Table 1). Following quality filtering, datasets were imputed using the Haplotype Reference Consortium imputation panel r1.1 2016 through the Michigan imputation server with default settings with phasing using the EAGLE v2.3 option ^23^. Genotypes were filtered for imputation quality R^2^□>□0.8, with the exception of rs10847864 (*HIP1R* GWAS signal) which was included as an independent signal on chromosome 12 (located ∼82mb upstream of *LRRK2*) despite slightly lower imputation quality in the Myers-Faroud (MF) ^24^ and Spanish Parkinson’s (IPDGC) cohorts (SPAIN3 and SPAIN4) (Supplementary Table 2). Principal components (PCs) were calculated as described in the Supplemental Methods.

**Table 1.**
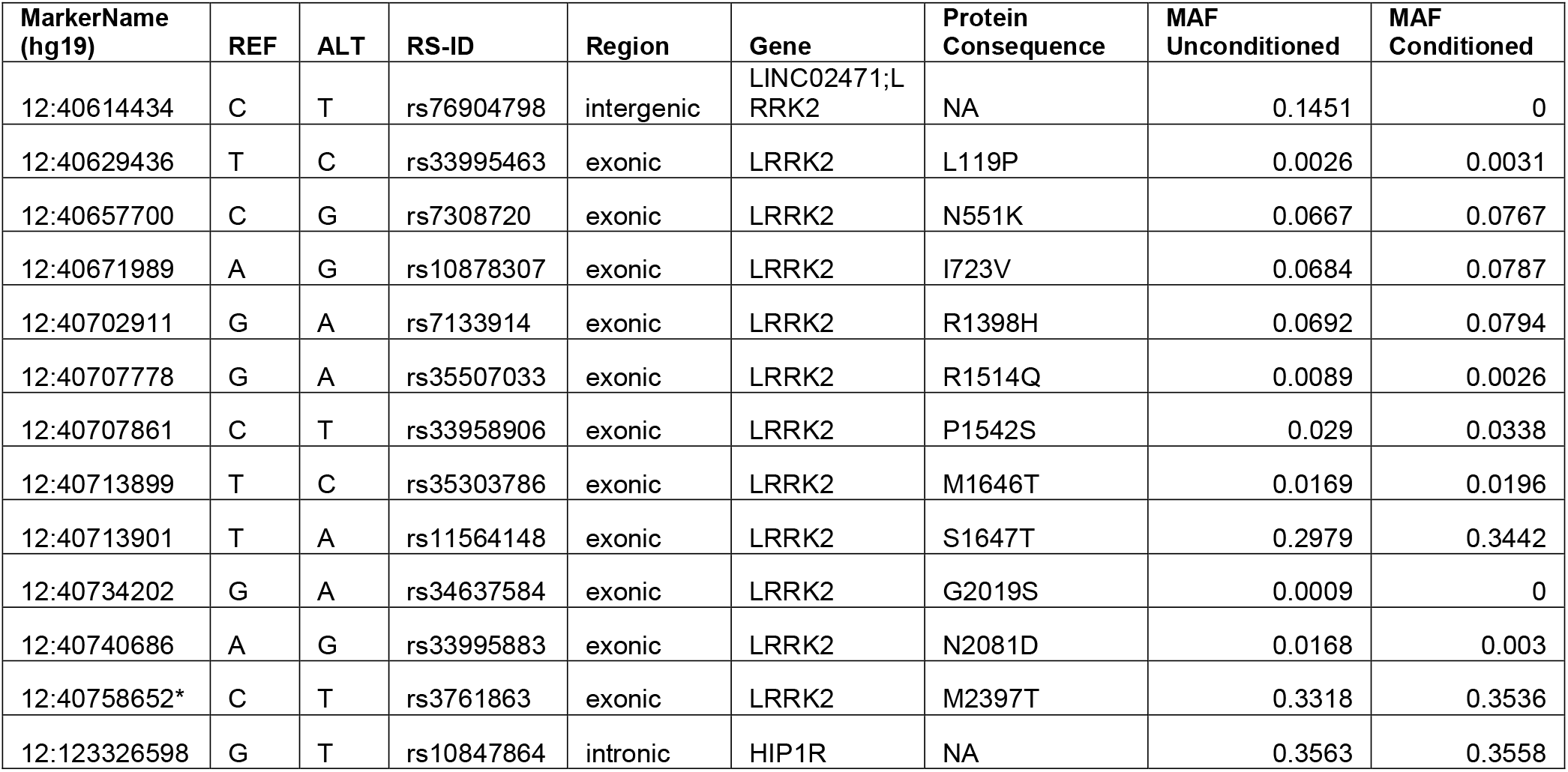
*LRRK2* variants examined in this study. All LRRK2 missense variants with frequency > 0.001 in the IPDGC, UK Biobank case-control and proxy-control datasets, in addition to rs76904798, p.G2019S, and the independent chromosome 12 signal in HIP1R (rs10847864). MarkerName denotes the chromosome and base pair position with respect to reference assembly GRCh37/hg19. REF and ALT denote the reference and alternate alleles used in the association analysis. MAF Unconditioned and MAF Conditioned denote the minor allele frequency from the combined IPDGC and UK Biobank datasets used in the unconditioned and conditioned (Δ p.G2019S Δ rs76904798) analyses, respectively.

| MarkerName (hg19) | REF | ALT | RS-ID | Region | Gene | Protein Consequence | MAF Unconditioned | MAF Conditioned |
| --- | --- | --- | --- | --- | --- | --- | --- | --- |
| 12:40614434 | C | T | rs76904798 | intergenic | LINC02471;LRRK2 | NA | 0.1451 | 0 |
| 12:40629436 | T | C | rs33995463 | exonic | LRRK2 | L119P | 0.0026 | 0.0031 |
| 12:40657700 | C | G | rs7308720 | exonic | LRRK2 | N551K | 0.0667 | 0.0767 |
| 12:40671989 | A | G | rs10878307 | exonic | LRRK2 | I723V | 0.0684 | 0.0787 |
| 12:40702911 | G | A | rs7133914 | exonic | LRRK2 | R1398H | 0.0692 | 0.0794 |
| 12:40707778 | G | A | rs35507033 | exonic | LRRK2 | R1514Q | 0.0089 | 0.0026 |
| 12:40707861 | C | T | rs33958906 | exonic | LRRK2 | P1542S | 0.029 | 0.0338 |
| 12:40713899 | T | C | rs35303786 | exonic | LRRK2 | M1646T | 0.0169 | 0.0196 |
| 12:40713901 | T | A | rs11564148 | exonic | LRRK2 | S1647T | 0.2979 | 0.3442 |
| 12:40734202 | G | A | rs34637584 | exonic | LRRK2 | G2019S | 0.0009 | 0 |
| 12:40740686 | A | G | rs33995883 | exonic | LRRK2 | N2081D | 0.0168 | 0.003 |
| 12:40758652* | C | T | rs3761863 | exonic | LRRK2 | M2397T | 0.3318 | 0.3536 |
| 12:123326598 | G | T | rs10847864 | intronic | HIP1R | NA | 0.3563 | 0.3558 |

**Table 2.** Association results of *LRRK2* variants. Odds ratios, 95% confidence intervals and P-values from the IPDGC and UK Biobank meta-analyses are reported for all LRRK2 variants examined in this study and the independent chromosome 12 signal in HIP1R (rs10847864). Heterozygosity estimates are based on a meta-analysis of the unconditioned datasets.

| RS-ID | Protein<br>Conse-<br>quence | Uncond-<br>itioned,<br>OR (95 CI) | Uncond-<br>itioned,<br>P-value | $\Delta$ p.G2019S<br>$\Delta$ rs76904798,<br>OR (95 CI) | $\Delta$ p.G2019S<br>$\Delta$ rs76904798,<br>P-value | $\Delta$ p.G2019S<br>$\Delta$ p.N2081D,<br>OR (95 CI) | $\Delta$ p.G2019S<br>$\Delta$ p.N2081D,<br>P-value | Het-ISq | Het-PVal |
| --- | --- | --- | --- | --- | --- | --- | --- | --- | --- |
| rs76904798 | NA | 1.12<br>(1.08-1.16) | 4.01E-09 | NA | NA | 1.11<br>(1.07-1.16) | 1.40E-07 | 23.793 | 0.04853 |
| rs33995463 | L119P | 0.94<br>(0.69-1.29) | 0.7223 | 0.91<br>(0.64-1.29) | 0.5976 | 0.91<br>(0.66-1.25) | 0.5521 | 5.687 | 0.7708 |
| rs7308720 | N551K | 0.9<br>(0.85-0.95) | 1.13E-04 | 0.93<br>(0.88-0.99) | 0.01506 | 0.91<br>(0.86-0.96) | 4.55E-04 | 8.957 | 0.8338 |
| rs10878307 | I723V | 1<br>(0.95-1.06) | 0.8648 | 1.03<br>(0.97-1.09) | 0.3545 | 1.03<br>(0.98-1.08) | 0.3042 | 23.402 | 0.05403 |
| rs7133914 | R1398H | 0.9<br>(0.86-0.95) | 1.31E-04 | 0.93<br>(0.87-0.98) | 0.01029 | 0.91<br>(0.86-0.96) | 6.92E-04 | 7.938 | 0.8925 |
| rs35507033 | R1514<br>Q | 1.13<br>(0.98-1.31) | 0.1002 | 0.98<br>(0.66-1.47) | 0.9402 | 1.14<br>(0.98-1.32) | 0.08066 | 9.926 | 0.7676 |
| rs33958906 | P1542S | 0.93<br>(0.86-1.01) | 0.0807 | 0.97<br>(0.88-1.05) | 0.4304 | 0.96<br>(0.88-1.04) | 0.2771 | 17.997 | 0.2069 |
| rs35303786 | M1646T | 1.18<br>(1.07-1.3) | 9.15E-04 | 1.16<br>(1.04-1.3) | 6.59E-03 | 1.19<br>(1.08-1.32) | 6.32E-04 | 21.122 | 0.09855 |
| rs11564148 | S1647T | 0.99<br>(0.96-1.02) | 0.4799 | 0.99<br>(0.96-1.03) | 0.7366 | 0.98<br>(0.95-1.01) | 0.1157 | 10.287 | 0.7409 |
| rs34637584 | G2019S | 9.02<br>(6.14-13.25) | 3.66E-29 | NA | NA | NA | NA | 4.34 | 0.993 |
| rs33995883 | N2081D | 1.18<br>(1.07-1.29) | 6.59E-04 | 1.2<br>(0.9-1.61) | 0.2054 | NA | NA | 17.291 | 0.241 |
| rs3761863 | M2397T | 1.01<br>(0.98-1.04) | 0.5904 | 1.04<br>(1-1.07) | 0.03988 | 1.01<br>(0.98-1.04) | 0.7049 | 20.418 | 0.1175 |
| rs10847864 | NA | 1.1<br>(1.07-1.13) | 8.76E-11 | 1.09 (1.06-1.13) | 1.65E-07 | 1.1 (1.06-1.13) | 3.42E-10 | 32.267 | 3.67E-03 |

#### UK Biobank data

The UK Biobank is a large study of approximately 500,000 individuals from the United Kingdom with a variety of phenotypic information, including information on PD status such as ICD-10 designation and self-report ^25^. PD cases were identified using field code 42033 and proxy cases were included based on their paternal PD status (data field 20107) and maternal PD status (data field 20110). We have previously shown that these proxy cases share genetic risk with PD cases ^15^. Genotype data was obtained as previously described ^15^ and was split into case-control and proxy-control datasets for separate GWAS. Quality control parameters can be found in the Supplemental Methods. After quality control, the case-control dataset consisted of individual-level genotypes from 1529 cases and 15,279 healthy controls, with the controls selected randomly from the pool of non-affected, non-proxy individuals in the biobank. The proxy-control dataset consisted of 13,404 proxy-cases and 140,655 healthy controls (Supplementary Table 1). Principal components (PCs) were calculated as described in the Supplemental Methods.

### Association analyses

Summary statistics were generated for each study using a logistic regression model on imputed genotypes from chromosome 12, followed by Firth regression when logistic regression failed to converge. IPDGC data were adjusted for biological sex, age and the first 5 PCs representing population substructure. Age at onset was used for PD cases and age at study for healthy controls, except in the Vance (dbGap phs000394) and Myers-Faroud ^24^ cohorts due to missing data. The UK Biobank data were adjusted for biological sex, age at recruitment, Townsend and the first five PCs. Genome-wide association study by proxy (GWAX) was carried out as previously described on UK Biobank proxy-case data ^15^ since these individuals are at-risk but not true cases. A fixed-effects model was fitted using METAL v2018-08-28 under default settings ^26^ to combine summary statistics across the 13 cohorts from IPDGC and the UK Biobank case-control and proxy-control datasets. After post GWAS filtering of multi-allelic variants, regional plots of the meta-analysis results were made around the *LRRK2* gene using LocusZoom v1.3 ^27^. Forest plots were generated using the metafor package in R. A summary of the association results for the *LRRK2* variants examined in this study are presented in Table 2 and Supplementary Table 3. We included an independent association signal on chromosome 12 at the *HIP1R* locus (rs10847864) identified in a previous PD meta-analysis ^15^. Statistical power was calculated as described in the Supplemental Methods.

### Co-inheritance analysis

We used two methods to compare the allelic distributions of the SNPs described in Table 1 before and after removing carriers of p.G2019S, rs76904798, p.N2081D, or a combination of these variants. First, we aggregated all samples used in the meta-analysis and calculated the percentage of carriers of a particular SNP that were excluded in the conditioned datasets (Supplementary Table 4). This was used as a disease-independent measure of the co-inheritance of the two variants. Second, we performed Fisher’s exact test in R with Bonferroni correction to assess whether there were significant differences in the allelic distributions of each SNP between the unconditioned and conditioned datasets. For this comparison, only cases and proxy-cases were included due to the large imbalance between case and control numbers in the UK Biobank. This analysis was performed separately for the IPDGC case, UK Biobank case and proxy-case datasets (Supplementary Table 5).

### Code availability

All of the code used in this study can be found at https://github.com/neurogenetics/LRRK2_conditional_v3.

## Results

### Included Data Overview

We included a total of 15 datasets, including 13 case-control cohorts (n_case=16,309, n_control=17,705), case-control data from the UK Biobank (n_case=1,529, n_control=15,279) and proxy-control data from the UK Biobank (n_proxy-case=13,404, n_control=140,655). Together, these sample sets comprised a total of 17,838 PD cases, 13,404 proxy-cases and 173,639 healthy controls of European ancestry. The mean age of onset for PD in the IPDGC cohorts, age at recruitment for the UK Biobank, and percentage female participants for each dataset are reported in Supplementary Table 1, where this information was available.

### *LRRK2* rs76904798 is independently associated with Parkinson’s disease risk from *LRRK2* coding variation

In the examination of the IPDGC, UK Biobank case-control and proxy-control datasets, we identified 10 nonsynonymous *LRRK2* coding variants with a minor allele frequency > 0.001 in all three datasets, as presented in Table 1. We verified the presence of these variants in the IPDGC datasets using whole□genome sequencing data ^28^ and in the UK Biobank datasets using whole□exome sequencing data ^29^. The concordance rates were very high (avg = 99.24%), showing that the imputation of these variants was highly accurate. The imputation quality and concordance values are presented in Supplementary Tables 2 and 6, respectively.

Four of these variants (p.N551K, p.R1398H, p.M1646T and p.N2081D) have been previously associated with either decreased or increased PD risk in Europeans ^7,8,30^, and the haplotype p.S1647T-p.M2397T has been suggested to confer a protective effect in Taiwanese individuals ^31^. Although none of the four previously associated variants reached genome-wide significance in our meta-analysis (Table 2, Supplementary Figures 1-4), the direction of effects were consistent with previous studies ^7,15^. We did not find any evidence of an association with PD for either p.S1647T or p.M2397T in this study (Supplementary Figures 5 and 6).

To confirm the effect of previously identified PD GWAS loci in *LRRK2*, we first verified the associations of the known pathogenic variant p.G2019S (OR: 9.02, 95% CI: 6.14-13.25, P=3.66E-29) and the 5’ non-coding variant rs76904798 (OR: 1.12, 95% CI: 1.08-1.16, P=4.01E-09) in a meta-analysis of all fifteen datasets (Table 2, Supplementary Figure 7). We then confirmed the independent association of rs76904798 with PD risk from p.G2019S and p.N2081D (Δ p.G2019S Δ p.N2081D, OR: 1.11, 95% CI: 1.07-1.16, P=1.40e−07) (Table 2, Supplementary Figure 8), the PD-linked *LRRK2* coding variants (Δ p.G2019S Δ p.N551K Δ p.R1398H Δ p.M1646T Δ p.N2081D, OR: 1.11, 95% CI: 1.06-1.16, P=2.144e−06), and all of the relatively rare *LRRK2* coding variants examined in this study (Δ p.G2019S Δ p.N551K Δ p.R1398H Δ p.M1646T Δ p.N2081D Δ p.L119P Δ p.I723V Δ p.R1514Q Δ p.P1542S, OR: 1.09, 95% CI: 1.04-1.15, P=0.0002082) (Figure 1). We used a Bonferroni-corrected P-value of P=0.006 to determine the independence of rs76904798, which is based on the number of *LRRK2* coding variants excluding p.G2019S that were examined in this study (Table 1), two of which (p.N551K-p.R1398H) represent the same allele (0.05/9=0.006). We also confirmed the independent association of rs76904798 by including the allele counts of the relatively rare *LRRK2* coding variants as covariates in a logistic regression model (OR: 1.11, 95% CI: 1.06-1.15, P=9.90e-07) (Supplementary Table 7).

**Figure 1.**
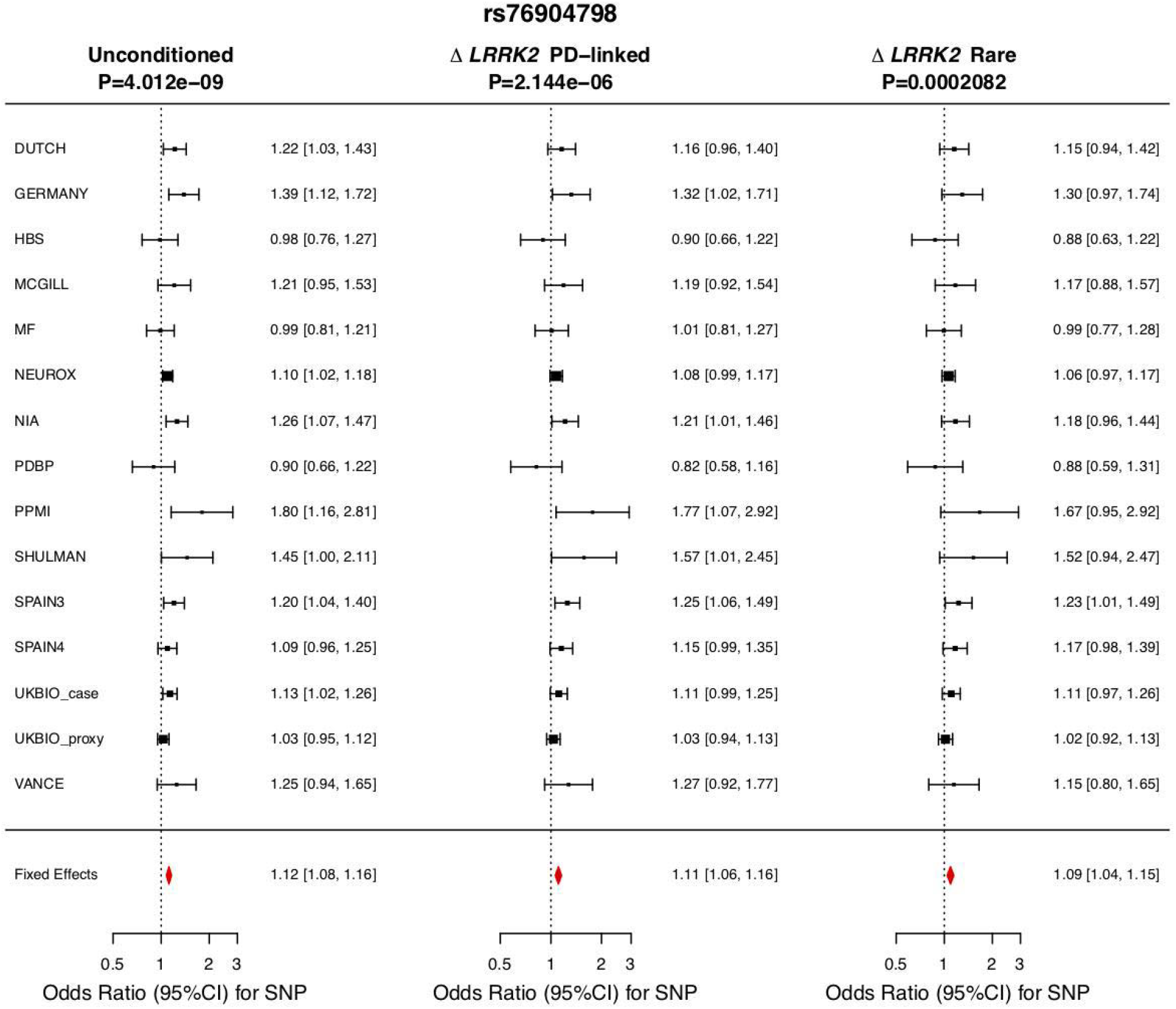
Meta-analysis of rs76904798 in the included datasets excluding (from left to right) 1) no samples 2) carriers of *LRRK2* p.G2019S, p.N551K, p.R1398H, p.M1646T and p.N2081D, and 3) carriers of *LRRK2* p.G2019S, p.N551K, p.R1398H, p.M1646T, p.N2081D, p.L119P, p.I723V, p.R1514Q and p.P1542S.

There were no other independent signals in the *LRRK2* region that reached genome-wide significance (P < 5e-8). The association of the 3’ intergenic variant rs190807041 disappears after conditioning on p.G2019S, and the 5’ signal driven by non-coding *LRRK2* variation explains the associations of both rs76904798 and the proximal intronic variant rs1491942 with PD risk (Figure 2).

**Figure 2.**
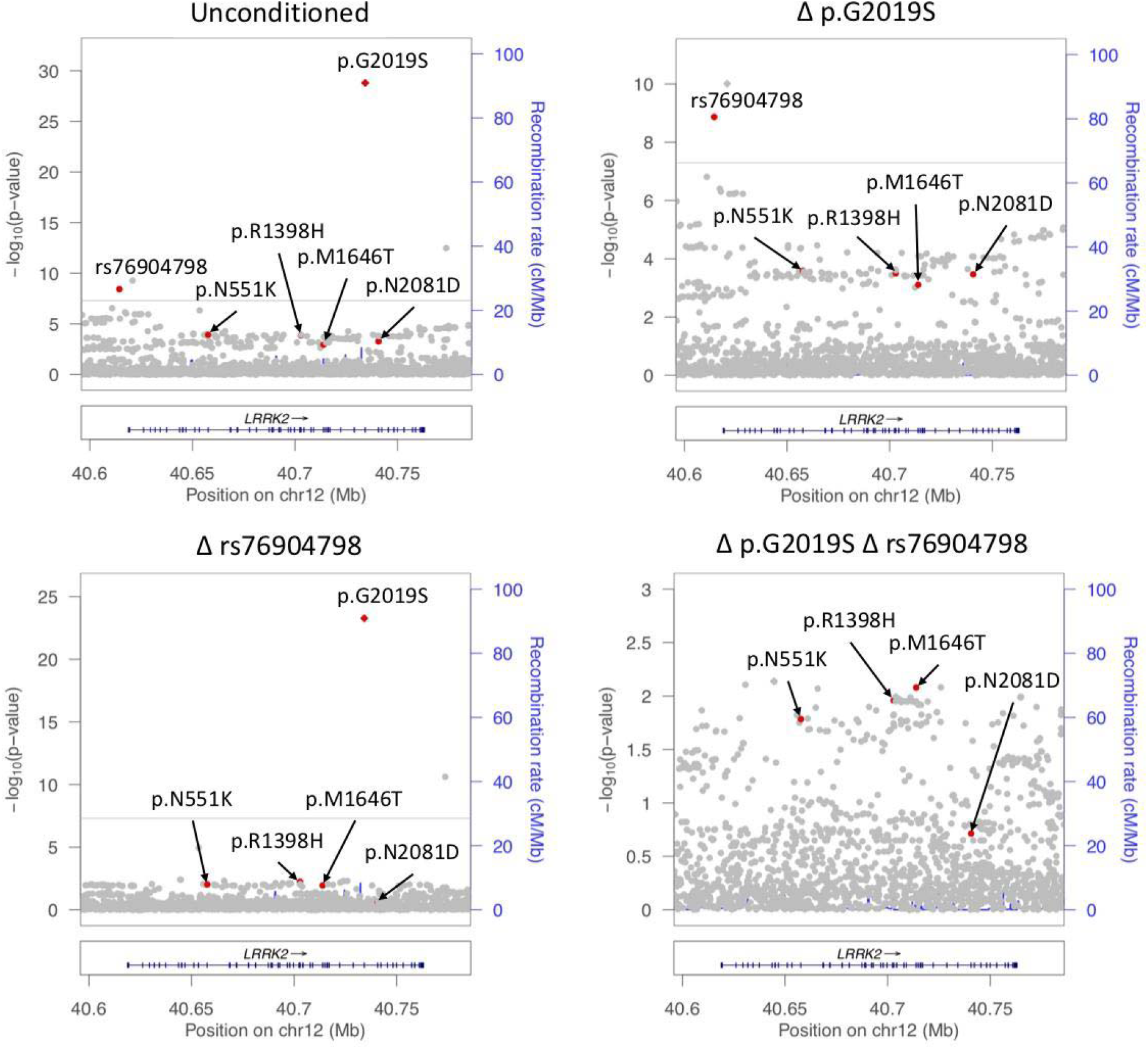
LocusZoom plot of *LRRK2* association with Parkinson’s disease risk. The top left panel shows the association signal at the *LRRK2* locus in the IPDGC and UK Biobank meta-analysis. The top right, bottom left and bottom right panels condition on p.G2019S, rs76904798, and both p.G2019S and rs76904798, respectively. The *LRRK2* variants p.N551K, p.R1398H, p.M1646T, p.N2081D, p.G2019S and rs76904798 are indicated by red dots unless the specified variant(s) were used in the conditional analysis.

### Conditional analysis of *LRRK2* exonic variants does not preclude an independent association of p.N551K-p.R1398H and p.M1646T with altered PD risk

To better understand the role of coding variation at the *LRRK2* locus in PD, we performed a meta-analysis excluding carriers of p.G2019S and rs76904798, representing the two independent GWAS signals at the locus. In total, there were 12,504 PD cases (total removed: 29.9%, p.G2019S carriers: 1.4%, rs76904798 carriers: 28.8%), 9,679 proxy-cases (total removed: 27.8%, p.G2019S carriers: 0.2%, rs76904798 carriers: 27.6%) and 127,254 controls (total removed: 26.7%, p.G2019S carriers: 0.1%, rs76904798 carriers: 26.6%) in the conditional analysis. After removing carriers of p.G2019S and rs76904798, there were no *LRRK2* variants that reached genome-wide significance. The results of the association analyses for the *LRRK2* variants examined in this study are presented in Table 2 and Supplementary Table 3, and forest plots are presented in and Supplementary Figures 1-6 and 8-12. Forest plots of *LRRK2* p.K1423K, which is in high LD with p.N551K-p.R1398H, and rs10847864 (*HIP1R* GWAS signal) are presented in Supplementary Figures 13 and 14, respectively. The association results for all *LRRK2* missense variants are presented in Supplementary Table 8.

With this sample size, we had ∼80% power to detect genotype relative risks ≥ 1.16 and ≥ 1.44 based on the minor allele frequencies (MAF) of p.M1646T and p.N2081D, respectively, in the conditional analysis (Table 1, Supplementary Figures 15 and 16). Likewise, we had ∼80% power to detect genotype relative risks ≤ 0.93 based on the MAF of p.N551K and p.R1398H in the conditional analysis (Table 1, Supplementary Figures 17 and 18). This calculation is based on the Bonferroni-corrected significance threshold of P=0.006.

None of the *LRRK2* variants examined in this study passed Bonferroni multiple test correction after excluding carriers of p.G2019S and rs76904798, including the previously PD-linked variants p.N551K (OR_condi: 0.93, 95% CI: 0.88-0.99, P=0.01506), p.R1398H (OR_condi: 0.93, 95% CI: 0.87-0.98, P=0.01029), p.M1646T (OR_condi: 1.16, 95% CI: 1.04-1.3, P=0.00659) and p.N2081D (OR_condi: 1.2, 95% CI: 0.91-1.61, P=0.2054) (Table 2, Supplementary Figures 1-4). LocusZoom plots of the stepwise conditional analysis are presented in Figure 2. However, p.N551K, p.R1398H and p.M1646T retained Bonferroni-corrected significance when p.G2019S and rs76904798 allele counts were included as covariates in a logistic regression model, though they still did not meet genome-wide significance (Supplementary Table 7). We also performed a meta-analysis excluding carriers of p.N2081D and p.G2019S since rs76904798 is often co-inherited with p.N2081D (Supplementary Figure 19). After solely removing p.G2019S carriers, *LRRK2* p.N551K, p.R1398H, p.M1646T and p.N2081D all passed Bonferroni multiple test correction (Supplementary Table 3).

### Rare *LRRK2* missense variants are co-inherited with rs76904798

To further investigate the effects of excluding rs76904798 carriers on the allele distribution of *LRRK2* coding variants, we calculated the percentage of carriers that were removed after excluding carriers of p.G2019S and rs76904798 in our datasets, as shown in Supplementary Table 4. We found that 86.64% of p.N2081D carriers and 79.07% of p.R1514Q carriers were excluded after removing carriers of rs76904798 from all 15 datasets. Only 0.19% of p.N2081D carriers and 0.08% of p.R1514Q carriers were excluded after removing p.G2019S carriers, all of which also carried rs76904798. Conversely, only 10.72% of rs76904798 carriers were excluded after removing p.N2081D carriers. Since the 5’ variant rs76904798 remained associated with PD after removing carriers of p.G2019S and p.N2081D (Supplementary Figure 8), the GWAS signal represented by rs76904798 is independent of p.N2081D and the previous significance reported for p.N2081D might be due to the rs76904798 signal.

We also found significant differences in the allele distributions of p.R1514Q, p.N2081D and p.S1647T after removing rs76904798 carriers from the IPDGC case, UK Biobank case, and proxy-case datasets (Supplementary Table 5). Although p.S1647T was significantly enriched among non-carriers of rs76904798, this variant was not associated with PD and around 17% of carriers were excluded after removing rs76904798 carriers. Only around 16% of p.N551K and p.R1398H carriers were excluded after removing rs76904798 carriers, and <1% after removing p.G2019S carriers. The complete results of the co-inheritance analysis are presented in Supplementary Tables 4 and 5, and the frequencies and allele distributions of the *LRRK2* variants examined in this study are presented in Supplementary Table 9.

## Discussion

It has been established that both rare and common variation at the *LRRK2* locus can influence PD susceptibility. Several large GWASes and meta-analyses have pointed to at least two independent association signals, represented by p.G2019S and 5’ non-coding variation, that reach genome-wide significance ^15^. However, the literature has often produced conflicting or inconclusive results when it comes to PD risk associated with common variation at the *LRRK2* locus. Interestingly, several *LRRK2* variants have been associated with other disorders, suggesting that there are shared pathological mechanisms that result from genetic changes in *LRRK2*. For example, *LRRK2* p.N2081D has been implicated as a shared risk variant in Crohn’s disease ^8^ and independent *LRRK2* ‘5 variation has been shown to be a modifier of progressive supranuclear palsy phenotypes ^32^. To refine our understanding of the pattern of *LRRK2* association with PD, we investigated the PD risk associated with *LRRK2* missense variants in a large meta-analysis of European-ancestry individuals.

First, we confirmed the genome-wide associations of p.G2019S and rs76904798 and identified potential associations of four other PD-linked *LRRK2* missense variants (p.N551K, p.R1398H, p.M1646T and p.N2081D), all of which passed multiple test correction. After removing carriers of the common non-coding variant rs76904798, we found that these missense variants do not pass Bonferroni correction, whereas they retained Bonferroni-corrected significance when rs76904798 was included as a covariate in a logistic regression model. These data therefore do not exclude the possibility that the putative protective haplotype p.N551K-p.R1398H and proposed risk factor p.M1646T are independently associated with PD from rs76904798. These variants, however, did not meet genome-wide significance in our study despite including a very large sample size.

In a recent analysis of whole-genome sequencing data from the Accelerating Medicines Partnership-Parkinson’s Disease (AMP-PD) cohort, Bryant et al found that the 5’ variant rs76904798 was associated with *LRRK2* p.R1514Q and p.N2081D ^33^. We confirmed the co-inheritance of these variants with rs76904798 in a dataset that is ∼20 times larger and also found a significant difference in the allele distribution of p.S1647T after removing rs76904798 carriers. Our results suggest that any association between p.N2081D and PD is very likely due to linkage disequilibrium with rs76904798. Bryant et al suggested that p.N2081D may be important in mediating disease risk associated with the 5’ non-coding variation. However, we establish here that rs76904798 is independently associated with PD from p.N2081D and therefore the role of p.N2081D in PD pathogenesis needs to be explored further and potentially reclassified.

Several *LRRK2* variants have been implicated in mechanisms that are relevant to PD pathogenesis. Previous studies of the p.G2019S mutation have shown that increased *LRRK2* kinase activity, typically estimated to be ∼2 fold compared to wild-type, has negative effects on neuronal survival ^34–37^. Both p.N2081D and p.M1646T are also associated with increased kinase activity compared to wild-type *LRRK2* ^8,38^, although the effect size is modest compared to p.G2019S. Despite this evidence, it is possible that prior data using transfection of plasmids in HEK293T cells are unable to disambiguate the effects of p.N2081D and rs76904798 as they are often present on the same haplotype. *LRRK2* p.M1646T has also been associated with increased glucocerebrosidase activity with a larger effect than p.G2019S ^30^. Additionally, *LRRK2* p.R1398H has been shown to affect GTPase and Wnt signaling activity in the opposite manner of pathogenic *LRRK2* mutations ^8,39–41^. A recent study has shown that both p.R1398H and p.N551K were able to counteract the putative pathogenic effects of p.G2019S in *Drosophila* models ^42^. The literature therefore suggests that each of these variants have measurable effects on protein function in cells and in vivo that are consistent with proposed direction of effects for risk vs protection. However, the relation between the functional impact and risk for PD needs to be further investigated for these coding variants.

Previous studies have also implicated the common *LRRK2* variant p.M2397T in several disorders. Although this variant does not appear to affect kinase activity ^36^, the allele M2397 has been reported to lower LRRK2 abundance due to protein destabilization ^43^, which has been linked to enhanced inflammatory responses in both Crohn’s disease ^43,44^ and leprosy ^45,46^. Despite the fact that p.M2397T has demonstrated a protective association with PD in a study consisting of 573 Taiwanese cases ^31^, and with MSA–a disease which shares many clinical and pathological features with PD–in a study consisting of 177 neuropathologically confirmed cases from America and the UK ^47^, we did not find any evidence that p.M2397T was associated with PD in this study. It is possible that previous studies linking *LRRK2* p.M2397T with PD and MSA were underpowered due to relatively small sample size, and further investigation in larger and diverse cohorts is warranted.

We note several limitations with this study. First, our use of imputed genotype data limits the accuracy of some genotypes, though very high concordance of the *LRRK2* coding variants with whole-genome and whole-exome sequence data shows that the imputation of these variants was highly accurate. Second, although we included a large amount of data in our meta-analysis, removing over one quarter of the samples in the conditional analysis limited our statistical power to detect less common small effect (OR ∼1.2) variants. Therefore, we cannot exclude that relatively common coding variants have a very small effect resulting in increased or decreased risk for PD. Further studies exploring the cumulative effects of common small effect variants could enhance our understanding of their genetic burden and should take into account the complex linkage disequilibrium structure of the *LRRK2* region. Third, our analysis was limited to individuals of European ancestry, so the results of this study may not extend beyond European-ancestry populations. In depth studies of PD-linked genes in diverse cohorts will help clarify the associations discussed here as well as broader disease mechanisms.

In conclusion, here we provide insights into the relationship between coding and non-coding variation at the *LRRK2* locus and demonstrate that the 5’ *LRRK2* non-coding GWAS signal represented by rs76904798 is independently associated with PD risk from *LRRK2* coding variation. These coding variants are therefore unlikely to drive the *LRRK2* GWAS signal in individuals of European ancestry and require additional genetic and functional studies to clarify their potential impact on disease state. Characterizing the PD susceptibility associated with *LRRK2* genetic variation is critical for anticipating disease progression and response to *LRRK2*-targeted therapeutics.

## Data Availability

All summary statistics are provided in the Supplementary tables. UK Biobank data can be requested at https://www.ukbiobank.ac.uk. All code used has been made available on Github: https://github.com/neurogenetics/LRRK2_conditional_v3.

## Acknowledgement

We would like to thank all of the subjects who donated their time and biological samples to be a part of this study. We also would like to thank all members of the International Parkinson Disease Genomics Consortium (IPDGC). See for a complete overview of members, acknowledgements and funding http://pdgenetics.org/partners. This work was supported in part by the Intramural Research Programs of the National Institute of Neurological Disorders and Stroke (NINDS), the National Institute on Aging (NIA), and the National Institute of Environmental Health Sciences both part of the National Institutes of Health, Department of Health and Human Services; project numbers 1ZIA-NS003154, Z01-AG000949-02 and Z01-ES101986. In addition this work was supported by the Department of Defense (award W81XWH-09-2-0128), and The Michael J Fox Foundation for Parkinson’s Research. This work was supported by National Institutes of Health grants R01NS037167, R01CA141668, P50NS071674, American Parkinson Disease Association (APDA); Barnes Jewish Hospital Foundation; Greater St Louis Chapter of the APDA. The KORA (Cooperative Research in the Region of Augsburg) research platform was started and financed by the Forschungszentrum für Umwelt und Gesundheit, which is funded by the German Federal Ministry of Education, Science, Research, and Technology and by the State of Bavaria. This study was also funded by the German Federal Ministry of Education and Research (BMBF) under the funding code 031A430A, the EU Joint Programme - Neurodegenerative Diseases Research (JPND) project under the aegis of JPND -www.jpnd.eu-through Germany, BMBF, funding code 01ED1406 and iMed - the Helmholtz Initiative on Personalized Medicine. This study is funded by the German National Foundation grant (DFG SH599/6-1) (grant to M.S), Michael J Fox Foundation, and MSA Coalition, USA (to M.S). The French GWAS work was supported by the French National Agency of Research (ANR-08-MNP-012). This study was also funded by France-Parkinson Association, Fondation de France, the French program “Investissements d’avenir” funding (ANR-10-IAIHU-06) and a grant from Assistance Publique-Hôpitaux de Paris (PHRC, AOR-08010) for the French clinical data. This study utilized the high-performance computational capabilities of the Biowulf Linux cluster at the National Institutes of Health, Bethesda, Md. (http://biowulf.nih.gov), and DNA panels, samples, and clinical data from the National Institute of Neurological Disorders and Stroke Human Genetics Resource Center DNA and Cell Line Repository. People who contributed samples are acknowledged in descriptions of every panel on the repository website. We thank the French Parkinson’s Disease Genetics Study Group and the Drug Interaction with genes (DIGPD) study group: Y Agid, M Anheim, F Artaud, A-M Bonnet, C Bonnet, F Bourdain, J-P Brandel, C Brefel-Courbon, M Borg, A Brice, E Broussolle, F Cormier-Dequaire, J-C Corvol, P Damier, B Debilly, B Degos, P Derkinderen, A Destée, A Dürr, F Durif, A Elbaz, D Grabli, A Hartmann, S Klebe, P. Krack, J Kraemmer, S Leder, S Lesage, R Levy, E Lohmann, L Lacomblez, G Mangone, L-L Mariani, A-R Marques, M Martinez, V Mesnage, J Muellner, F Ory-Magne, F Pico, V Planté-Bordeneuve, P Pollak, O Rascol, K Tahiri, F Tison, C Tranchant, E Roze, M Tir, M Vérin, F Viallet, M Vidailhet, A You. We also thank the members of the French 3C Consortium: A Alpérovitch, C Berr, C Tzourio, and P Amouyel for allowing us to use part of the 3C cohort, and D Zelenika for support in generating the genome-wide molecular data. We thank P Tienari (Molecular Neurology Programme, Biomedicum, University of Helsinki), T Peuralinna (Department of Neurology, Helsinki University Central Hospital), L Myllykangas (Folkhalsan Institute of Genetics and Department of Pathology, University of Helsinki), and R Sulkava (Department of Public Health and General Practice Division of Geriatrics, University of Eastern Finland) for the Finnish controls (Vantaa85+ GWAS data). This study was also funded by the Sigrid Juselius Foundation (KM). We used genome-wide association data generated by the Wellcome Trust Case-Control Consortium 2 (WTCCC2) from UK patients with Parkinson’s disease and UK control individuals from the 1958 Birth Cohort and National Blood Service. UK population control data was made available through WTCCC1. This study was supported by the Medical Research Council and Wellcome Trust disease centre (grant WT089698/Z/09/Z to NW, JHa, and ASc). As with previous IPDGC efforts, this study makes use of data generated by the Wellcome Trust Case-Control Consortium. A full list of the investigators who contributed to the generation of the data is available from www.wtccc.org.uk. Funding for the project was provided by the Wellcome Trust under award 076113, 085475 and 090355. This study was also supported by Parkinson’s UK (grants 8047 and J-0804) and the Medical Research Council (G0700943 and G1100643). Sequencing and genotyping done in McGill University was supported by grants from the Michael J. Fox Foundation, the Canadian Consortium on Neurodegeneration in Aging (CCNA), the Canada First Research Excellence Fund (CFREF), awarded to McGill University for the Healthy Brains for Healthy Lives (HBHL) program and Parkinson’s Society Canada. The access to part of the participants at McGill has been made possible thanks to the Quebec Parkinson’s Network (http://rpq-qpn.ca/en/). DNA extraction work that was done in the UK was undertaken at University College London Hospitals, University College London, who received a proportion of funding from the Department of Health’s National Institute for Health Research Biomedical Research Centres funding. This study was supported in part by the Wellcome Trust/Medical Research Council Joint Call in Neurodegeneration award (WT089698) to the Parkinson’s Disease Consortium (UKPDC), whose members are from the UCL Institute of Neurology, University of Sheffield, and the Medical Research Council Protein Phosphorylation Unit at the University of Dundee. We thank the Quebec Parkinson’s Network (http://rpq-qpn.org) and its members. Harvard NeuroDiscovery Biomarker Study (HBS) is a collaboration of HBS investigators and funded through philanthropy and NIH and Non-NIH funding sources. The HBS Investigators have not participated in reviewing the data analysis or content of the manuscript. PPMI – a public-private partnership – is funded by the Michael J. Fox Foundation for Parkinson’s Research and funding partners, the full names of all of the PPMI funding partners can be found at www.ppmi-info.org/fundingpartners. The PPMI Investigators have not participated in reviewing the data analysis or content of the manuscript. For up-to-date information on the study, visit www.ppmi-info.org. Parkinson’s Disease Biomarker Program (PDBP) consortium is supported by the National Institute of Neurological Disorders and Stroke (NINDS) at the National Institutes of Health. A full list of PDBP investigators can be found at https://pdbp.ninds.nih.gov/policy. The PDBP Investigators have not participated in reviewing the data analysis or content of the manuscript.

## Authors’ Roles

Concept and design: CB, HLL

Statistical analysis: JL, MAN, CB, HLL

Contributed expertise, data or DNA samples: XR, RL, MC, ZGO

Drafting of the manuscript: All

Critical revision of the manuscript: All

## References

1. Paisán-Ruíz C, Jain S, Evans EW, et al. Cloning of the gene containing mutations that cause PARK8-linked Parkinson’s disease. Neuron 2004;44(4):595–600.

2. Zimprich A, Biskup S, Leitner P, et al. Mutations in LRRK2 cause autosomal-dominant parkinsonism with pleomorphic pathology. Neuron 2004;44(4):601–607.

3. Healy DG, Falchi M, O’Sullivan SS, et al. Phenotype, genotype, and worldwide genetic penetrance of LRRK2-associated Parkinson’s disease: a case-control study. Lancet Neurol. 2008;7(7):583–590.

4. Kluss JH, Mamais A, Cookson MR. LRRK2 links genetic and sporadic Parkinson’s disease. Biochem. Soc. Trans. 2019;47(2):651.[cited 2021 Feb 9]

5. Bonet-Ponce L, Beilina A, Williamson CD, et al. LRRK2 mediates tubulation and vesicle sorting from lysosomes [Internet]. Science Advances 2020;6(46)[cited 2021 Feb 24] Available from: https://www.ncbi.nlm.nih.gov/pmc/articles/PMC7673727/

6. Tolosa E, Vila M, Klein C, Rascol O. LRRK2 in Parkinson disease: challenges of clinical trials. Nat. Rev. Neurol. 2020;16(2):97–107.

7. Ross OA, Soto-Ortolaza AI, Heckman MG, et al. Association of LRRK2 exonic variants with susceptibility to Parkinson’s disease: a case-control study. Lancet Neurol. 2011;10(10):898–908.

8. Hui KY, Fernandez-Hernandez H, Hu J, et al. Functional variants in the gene confer shared effects on risk for Crohn’s disease and Parkinson’s disease [Internet]. Sci. Transl. Med. 2018;10(423)Available from: http://dx.doi.org/10.1126/scitranslmed.aai7795

9. Wu X, Tang K-F, Li Y, et al. Quantitative assessment of the effect of LRRK2 exonic variants on the risk of Parkinson’s disease: A meta-analysis [Internet]. Parkinsonism & Related Disorders 2012;18(6):722–730.Available from: http://dx.doi.org/10.1016/j.parkreldis.2012.04.013

10. Tan EK, Peng R, Teo YY, et al. Multiple LRRK2 variants modulate risk of Parkinson disease: a Chinese multicenter study [Internet]. Hum. Mutat. 2010;31(5) [cited 2021 Mar 29] Available from: https://pubmed.ncbi.nlm.nih.gov/20186690/

11. Ross OA, Wu YR, Lee MC, et al. Analysis of Lrrk2 R1628P as a risk factor for Parkinson’s disease [Internet]. Ann. Neurol. 2008;64(1) [cited 2021 Mar 29] Available from: https://pubmed.ncbi.nlm.nih.gov/18412265/

12. Farrer MJ, Stone JT, Lin CH, et al. Lrrk2 G2385R is an ancestral risk factor for Parkinson’s disease in Asia [Internet]. Parkinsonism Relat. Disord. 2007;13(2) [cited 2021 Mar 29] Available from: https://pubmed.ncbi.nlm.nih.gov/17222580/

13. Di Fonzo A, Wu-Chou YH, Lu CS, et al. A common missense variant in the LRRK2 gene, Gly2385Arg, associated with Parkinson’s disease risk in Taiwan [Internet]. Neurogenetics 2006;7(3) [cited 2021 Mar 29] Available from: https://pubmed.ncbi.nlm.nih.gov/16633828/

14. González-Fernández MC, Lezcano E, Ross OA, et al. Lrrk2-associated parkinsonism is a major cause of disease in Northern Spain [Internet]. Parkinsonism Relat. Disord. 2007;13(8) [cited 2021 Mar 29] Available from: https://pubmed.ncbi.nlm.nih.gov/17540608/

15. Nalls MA, Blauwendraat C, Vallerga CL, et al. Identification of novel risk loci, causal insights, and heritable risk for Parkinson’s disease: a meta-analysis of genome-wide association studies. Lancet Neurol. 2019;18(12):1091–1102.[cited 2020 Nov 24]

16. Trabzuni D, Ryten M, Emmett W, et al. Fine-mapping, gene expression and splicing analysis of the disease associated LRRK2 locus. PLoS One 2013;8(8):e70724.

17. Heckman MG, Labbé C, Kolicheski AL, et al. Fine-mapping of the non-coding variation driving the Caucasian LRRK2 GWAS signal in Parkinson’s disease [Internet]. Parkinsonism Relat. Disord. 2021;83[cited 2021 Jan 19] Available from: https://pubmed.ncbi.nlm.nih.gov/33454605/

18. Soto-Ortolaza AI, Heckman MG, Labbé C, et al. GWAS risk factors in Parkinson’s disease: LRRK2 coding variation and genetic interaction with PARK16. Am. J. Neurodegener. Dis. 2013;2(4):287.[cited 2021 Jan 27]

19. Li YI, Wong G, Humphrey J, Raj T. Prioritizing Parkinson’s disease genes using population-scale transcriptomic data. Nat. Commun. 2019;10(1):1–10.[cited 2021 Jan 27]

20. Ryan KJ, White CC, Patel K, et al. Context-specific effects of neurodegenerative disease variants in a model of human microglia [Internet]. Sci. Transl. Med. 2017;9(421) [cited 2021 Feb 2] Available from: https://www.ncbi.nlm.nih.gov/pmc/articles/PMC5945290/

21. Langston RG, Beilina A, Reed X, et al. Association of a Common Genetic Variant with Parkinson’s Disease is Propagated through Microglia [Internet]. Cold Spring Harbor Laboratory 2021;2021.01.15.426824. [cited 2021 Feb 23] Available from: https://www.biorxiv.org/content/10.1101/2021.01.15.426824v1.abstract

22. Iwaki H, Blauwendraat C, Leonard HL, et al. Genetic risk of Parkinson disease and progression:: An analysis of 13 longitudinal cohorts [Internet]. Neurology: Genetics 2019;5(4) [cited 2021 Feb 2] Available from: https://www.ncbi.nlm.nih.gov/pmc/articles/PMC6659137/

23. Das S, Forer L, Schönherr S, et al. Next-generation genotype imputation service and methods. Nat. Genet. 2016;48(10):1284–1287.

24. Pankratz N, Beecham GW, DeStefano AL, et al. Meta-analysis of Parkinson’s disease: identification of a novel locus, RIT2 [Internet]. Ann. Neurol. 2012;71(3) [cited 2021 Feb 24] Available from: https://pubmed.ncbi.nlm.nih.gov/22451204/

25. Bycroft C, Freeman C, Petkova D, et al. The UK Biobank resource with deep phenotyping and genomic data. Nature 2018;562(7726):203–209.

26. Willer CJ, Li Y, Abecasis GR. METAL: fast and efficient meta-analysis of genomewide association scans. Bioinformatics 2010;26(17):2190–2191.

27. Pruim RJ, Welch RP, Sanna S, et al. LocusZoom: regional visualization of genome-wide association scan results. Bioinformatics 2010;26(18):2336–2337.

28. Iwaki H, Leonard HL, Makarious MB, et al. Accelerating Medicines Partnership: Parkinson’s Disease. Genetic Resource. medRxiv 2020;2020.11.19.20235192.[cited 2021 Feb 11]

29. Szustakowski JD, Balasubramanian S, Sasson A, et al. Advancing Human Genetics Research and Drug Discovery through Exome Sequencing of the UK Biobank. medRxiv 2020;2020.11.02.20222232.[cited 2021 Feb 11]

30. Sosero YL, Yu E, Krohn L, et al. LRRK2 p.M1646T is associated with glucocerebrosidase activity and with Parkinson’s disease [Internet]. [date unknown];Available from: http://dx.doi.org/10.1101/2020.09.23.20197558

31. Wu YR, Chang KH, Chang WT, et al. Genetic variants ofLRRK2 in Taiwanese Parkinson’s disease [Internet]. PLoS One 2013;8(12) [cited 2021 Feb 1] Available from: https://pubmed.ncbi.nlm.nih.gov/24339985/

32. Jabbari E, Koga S, Valentino RR, et al. Genetic determinants of survival in progressive supranuclear palsy: a genome-wide association study [Internet]. Lancet Neurol. 2021;20(2) [cited 2021 Feb 11] Available from: https://pubmed.ncbi.nlm.nih.gov/33341150/

33. Bryant N, Malpeli N, Ziaee J, et al. Identification of LRRK2 Missense Variants in the Accelerating Medicines Partnership Parkinson’s Disease Cohort [Internet]. Hum. Mol. Genet. 2021;[cited 2021 Mar 1] Available from: https://pubmed.ncbi.nlm.nih.gov/33640967/

34. Smith WW, Pei Z, Jiang H, et al. Kinase activity of mutant LRRK2 mediates neuronal toxicity. Nat. Neurosci. 2006;9(10):1231–1233.

35. Greggio E, Jain S, Kingsbury A, et al. Kinase activity is required for the toxic effects of mutant LRRK2/dardarin. Neurobiol. Dis. 2006;23(2):329–341.

36. West AB, Moore DJ, Biskup S, et al. Parkinson’s disease-associated mutations in leucine-rich repeat kinase 2 augment kinase activity [Internet]. Proc. Natl. Acad. Sci. U. S. A. 2005;102(46) [cited 2021 Jan 18] Available from: https://pubmed.ncbi.nlm.nih.gov/16269541/

37. Luzón-Toro B, de la Torre E R, Delgado A, et al. Mechanistic insight into the dominant mode of the Parkinson’s disease-associated G2019S LRRK2 mutation [Internet]. Hum. Mol. Genet. 2007;16(17) [cited 2021 Jan 18] Available from: https://pubmed.ncbi.nlm.nih.gov/17584768/

38. Refai FS, Ng SH, Tan E-K. Evaluating LRRK2 genetic variants with unclear pathogenicity. Biomed Res. Int. 2015;2015:678701.

39. Nixon-Abell J, Berwick DC, Grannó S, et al. Protective LRRK2 R1398H Variant Enhances GTPase and Wnt Signaling Activity. Front. Mol. Neurosci. 2016;9:18.

40. Berwick DC, Harvey K. LRRK2 functions as a Wnt signaling scaffold, bridging cytosolic proteins and membrane-localized LRP6 [Internet]. Hum. Mol. Genet. 2012;21(22) [cited 2021 Jan 21] Available from: https://pubmed.ncbi.nlm.nih.gov/22899650/

41. Berwick DC, Javaheri B, Wetzel A, et al. Pathogenic LRRK2 variants are gain-of-function mutations that enhance LRRK2-mediated repression of β-catenin signaling [Internet]. Mol. Neurodegener. 2017;12(1) [cited 2021 Feb 2] Available from: https://pubmed.ncbi.nlm.nih.gov/28103901/

42. Toh J, Chua LL, Ho P, et al. Identification of Targets from LRRK2 Rescue Phenotypes [Internet]. Cells 2021;10(1) [cited 2021 Feb 2] Available from: https://www.ncbi.nlm.nih.gov/pmc/articles/PMC7824855/

43. Liu Z, Lee J, Krummey S, et al. Leucine-rich repeat kinase 2 (LRRK2) regulates inflammatory bowel disease through the Nuclear Factor of Activated T cells (NFAT). Nat. Immunol. 2011;12(11):1063.[cited 2021 Feb 1]

44. Ikezu T, Koro L, Wolozin B, et al. Crohn’s and Parkinson’s Disease-Associated LRRK2 Mutations Alter Type II Interferon Responses in Human CD14 + Blood Monocytes Ex Vivo [Internet]. J. Neuroimmune Pharmacol. 2020;15(4) [cited 2021 Feb 1] Available from: https://pubmed.ncbi.nlm.nih.gov/32180132/

45. Wang D, Xu L, Lv L, et al. Association of the LRRK2 genetic polymorphisms with leprosy in Han Chinese from Southwest China [Internet]. Genes Immun. 2015;16(2) [cited 2021 Feb 1] Available from: https://pubmed.ncbi.nlm.nih.gov/25521227/

46. Fava VM, Manry J, Cobat A, et al. A Missense LRRK2 Variant Is a Risk Factor for Excessive Inflammatory Responses in Leprosy [Internet]. PLoS Negl. Trop. Dis. 2016;10(2) [cited 2021 Feb 1] Available from: https://www.ncbi.nlm.nih.gov/pmc/articles/PMC4742274/

47. Heckman MG, Schottlaender L, Soto-Ortolaza AI, et al. LRRK2 exonic variants and risk of multiple system atrophy [Internet]. Neurology 2014;83(24) [cited 2021 Jan 31] Available from: https://pubmed.ncbi.nlm.nih.gov/25378673/

