## Supplementary Figures for "*LRRK2* coding variants and the risk of Parkinson’s disease"

**Supplementary Figure 1.** Meta-analysis of p.N551K in the included datasets excluding (from left to right) 1) no samples 2) carriers of rs76904798 and p.G2019S and 3) carriers of p.N2081D and p.G2019S.

**Supplementary Figure 2.** Meta-analysis of p.R1398H in the included datasets excluding (from left to right) 1) no samples 2) carriers of rs76904798 and p.G2019S and 3) carriers of p.N2081D and p.G2019S.

**Supplementary Figure 3.** Meta-analysis of p.M1646T in the included datasets excluding (from left to right) 1) no samples 2) carriers of rs76904798 and p.G2019S and 3) carriers of p.N2081D and p.G2019S.

**Supplementary Figure 4.** Meta-analysis of p.N2081D in the included datasets excluding (from left to right) 1) no samples and 2) carriers of rs76904798 and p.G2019S.

**Supplementary Figure 5.** Meta-analysis of p.S1647T in the included datasets excluding (from left to right) 1) no samples 2) carriers of rs76904798 and p.G2019S and 3) carriers of p.N2081D and p.G2019S.

**Supplementary Figure 6.** Meta-analysis of p.M2397T in the included datasets excluding (from left to right) 1) no samples 2) carriers of rs76904798 and p.G2019S and 3) carriers of p.N2081D and p.G2019S.

**Supplementary Figure 7.** Meta-analysis of (from left to right) 1) p.G2019S and 2) rs76904798 in the included datasets.

**Supplementary Figure 8.** Meta-analysis of rs76904798 in the included datasets excluding (from left to right) 1) no samples and 2) carriers of p.N2081D and p.G2019S.

**Supplementary Figure 9.** Meta-analysis of p.L119P in the included datasets excluding (from left to right) 1) no samples 2) carriers of rs76904798 and p.G2019S and 3) carriers of p.N2081D and p.G2019S.

**Supplementary Figure 10.** Meta-analysis of p.I723V in the included datasets excluding (from left to right) 1) no samples 2) carriers of rs76904798 and p.G2019S and 3) carriers of p.N2081D and p.G2019S.

**Supplementary Figure 11.** Meta-analysis of p.R1514Q in the included datasets excluding (from left to right) 1) no samples 2) carriers of rs76904798 and p.G2019S and 3) carriers of p.N2081D and p.G2019S.

**Supplementary Figure 12.** Meta-analysis of p.P1542S in the included datasets excluding (from left to right) 1) no samples 2) carriers of rs76904798 and p.G2019S and 3) carriers of p.N2081D and p.G2019S.

**Supplementary Figure 13.** Meta-analysis of p.K1423K in the included datasets excluding (from left to right) 1) no samples 2) carriers of rs76904798 and p.G2019S and 3) carriers of p.N2081D and p.G2019S.

**Supplementary Figure 14.** Meta-analysis of rs10847864 in the included datasets excluding (from left to right) 1) no samples 2) carriers of rs76904798 and p.G2019S and 3) carriers of p.N2081D and p.G2019S.

**Supplementary Figure 15.** GAS Power Calculator interface for LRRK2 p.M1646T in the conditional analysis. The left panel shows the input parameters, the right panel shows the genotype relative risks detected at a given power level with these input parameters, and the bottom panel shows the results of the power calculation.

**Supplementary Figure 16.** GAS Power Calculator interface for LRRK2 p.N2081D in the conditional analysis. The left panel shows the input parameters, the right panel shows the genotype relative risks detected at a given power level with these input parameters, and the bottom panel shows the results of the power calculation.

**Supplementary Figure 17.** GAS Power Calculator interface for LRRK2 p.N551K in the conditional analysis. The left panel shows the input parameters, the right panel shows the genotype relative risks detected at a given power level with these input parameters, and the bottom panel shows the results of the power calculation. Since p.N551K has demonstrated a protective association with PD, the resulting relative risk was modified as follows: the natural logarithm was taken, multiplied by -1, and exponentiated.

**Supplementary Figure 18.** GAS Power Calculator interface for LRRK2 p.R1398H in the conditional analysis. The left panel shows the input parameters, the right panel shows the genotype relative risks detected at a given power level with these input parameters, and the bottom panel shows the results of the power calculation. Since p.R1398H has demonstrated a protective association with PD, the resulting relative risk was modified as follows: the natural logarithm was taken, multiplied by -1, and exponentiated.

**Supplementary Figure 19.** LocusZoom plot of *LRRK2* association with Parkinson’s disease risk conditioned on p.N2081D. The left panel shows the association signal at the *LRRK2* locus in the IPDGC and UK Biobank meta-analysis conditioned on p.N2081D, and the right panel conditions on both p.G2019S and p.N2081D. The LRRK2 variants p.N551K, p.R1398H, p.M1646T, p.G2019S and rs76904798 are indicated by red dots.

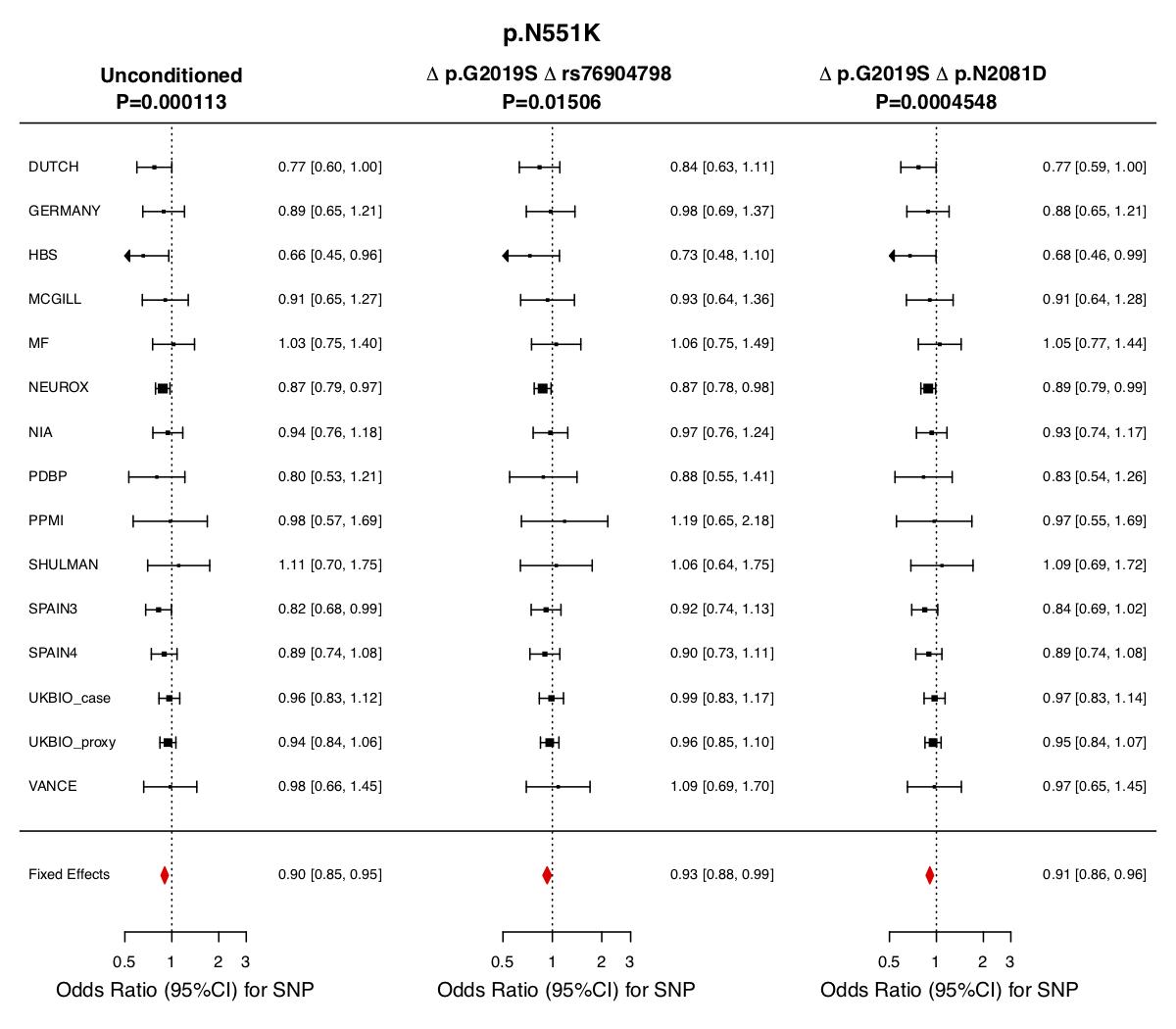

**Supplementary Figure 1.** Meta-analysis of p.N551K in the included datasets excluding (from left to right) 1) no samples 2) carriers of rs76904798 and p.G2019S and 3) carriers of p.N2081D and p.G2019S.

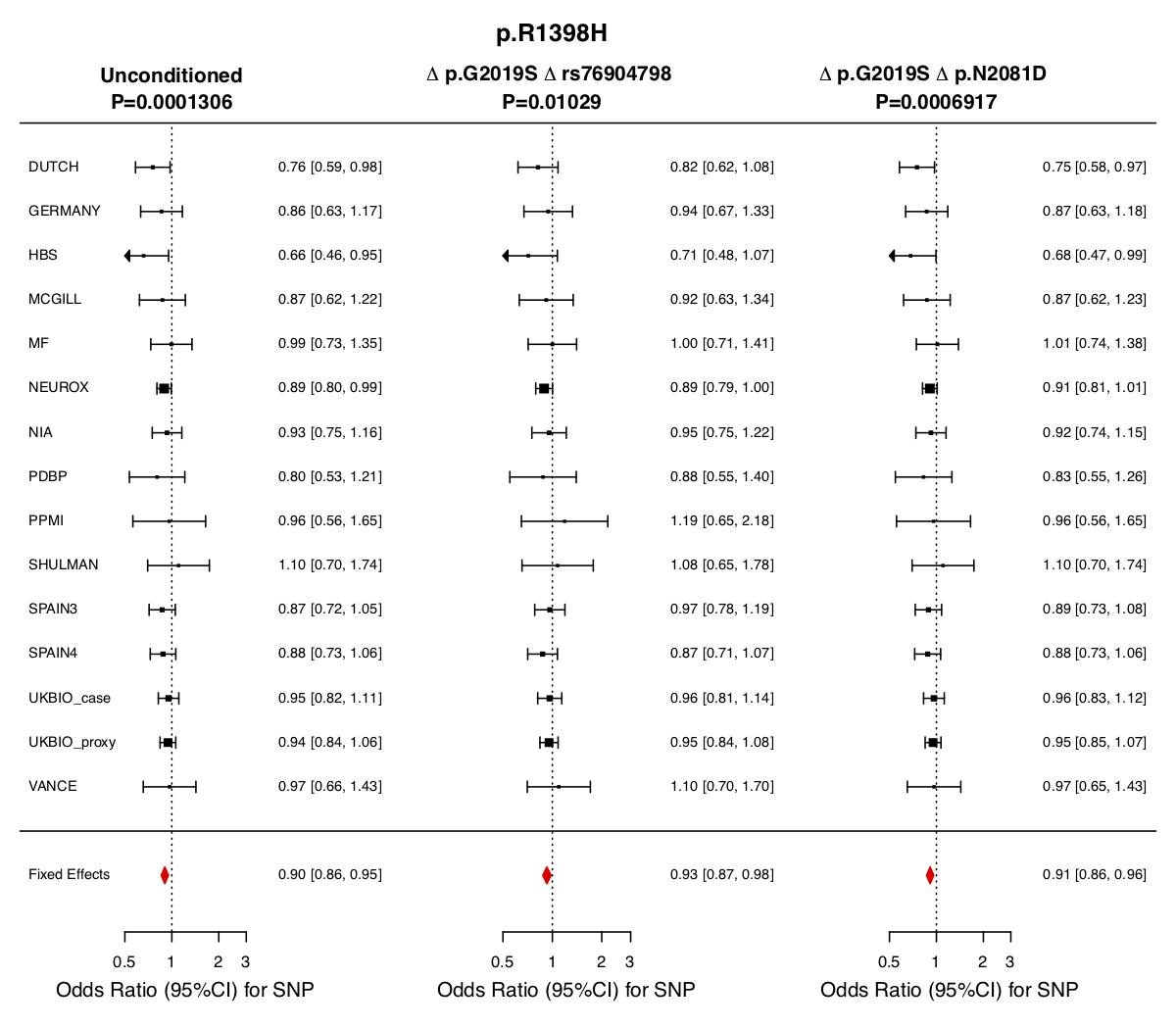

**Supplementary Figure 2.** Meta-analysis of p.R1398H in the included datasets excluding (from left to right) 1) no samples 2) carriers of rs76904798 and p.G2019S and 3) carriers of p.N2081D and p.G2019S.

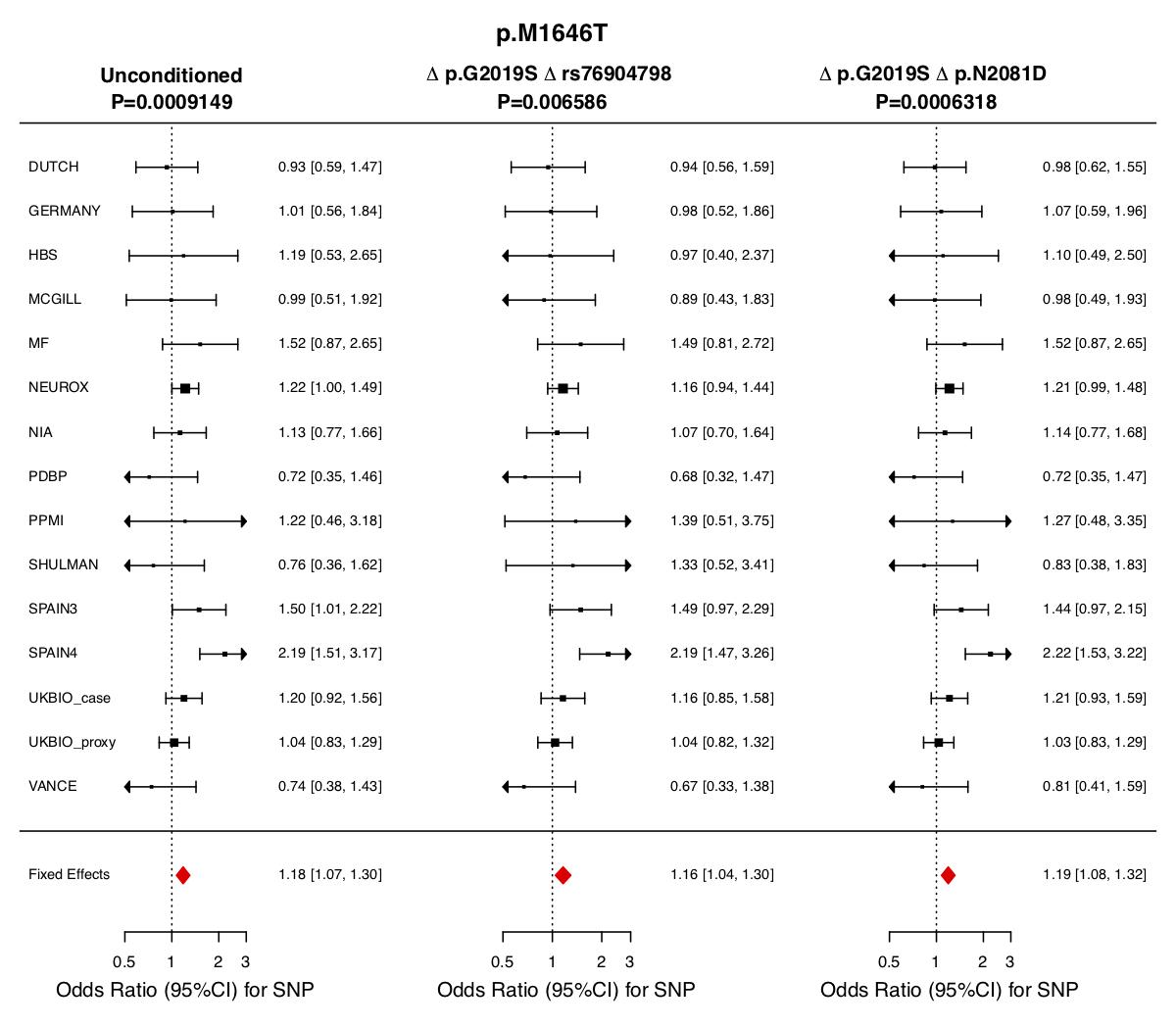

**Supplementary Figure 3.** Meta-analysis of p.M1646T in the included datasets excluding (from left to right) 1) no samples 2) carriers of rs76904798 and p.G2019S and 3) carriers of p.N2081D and p.G2019S.

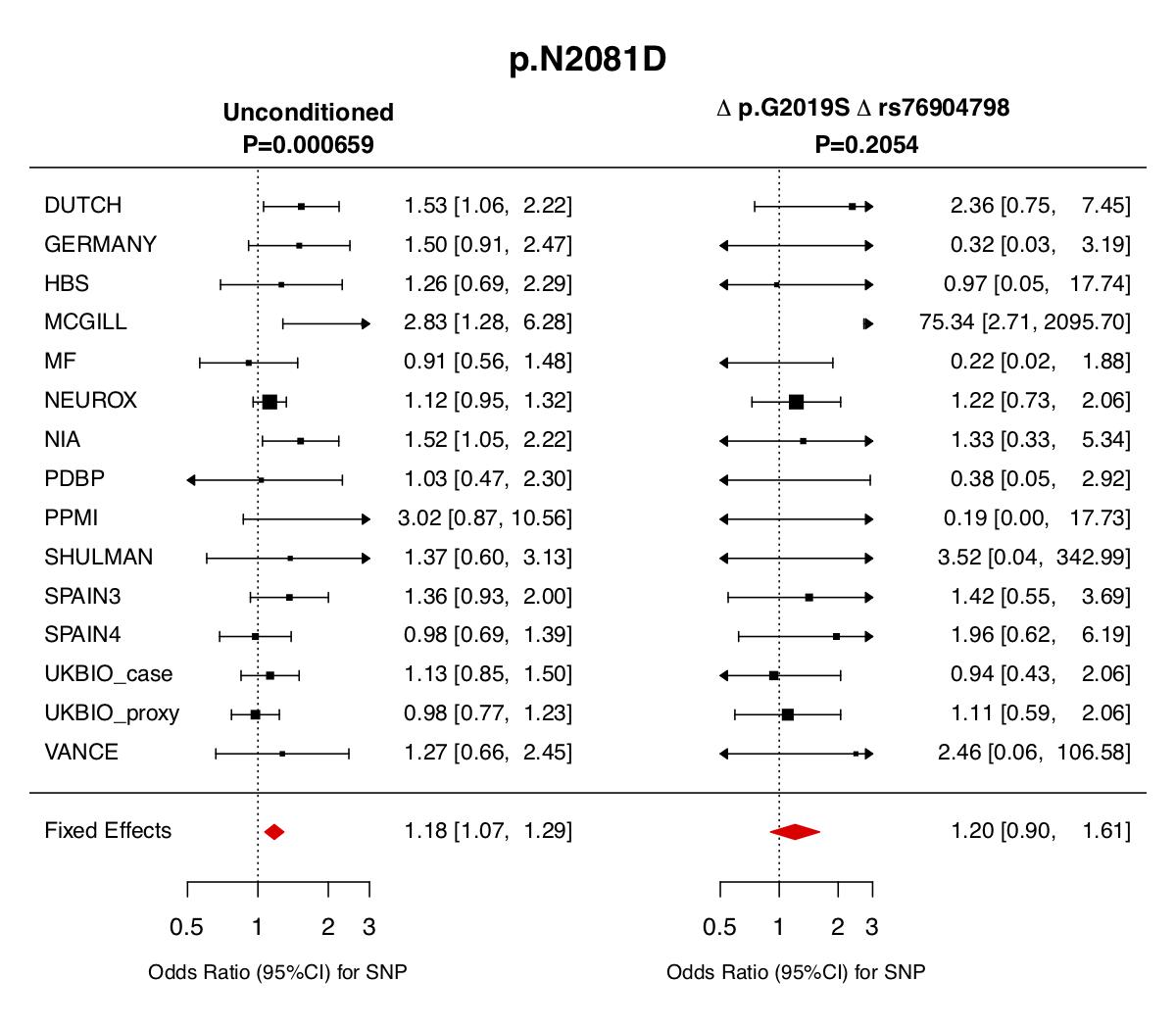

**Supplementary Figure 4.** Meta-analysis of p.N2081D in the included datasets excluding (from left to right) 1) no samples and 2) carriers of rs76904798 and p.G2019S.

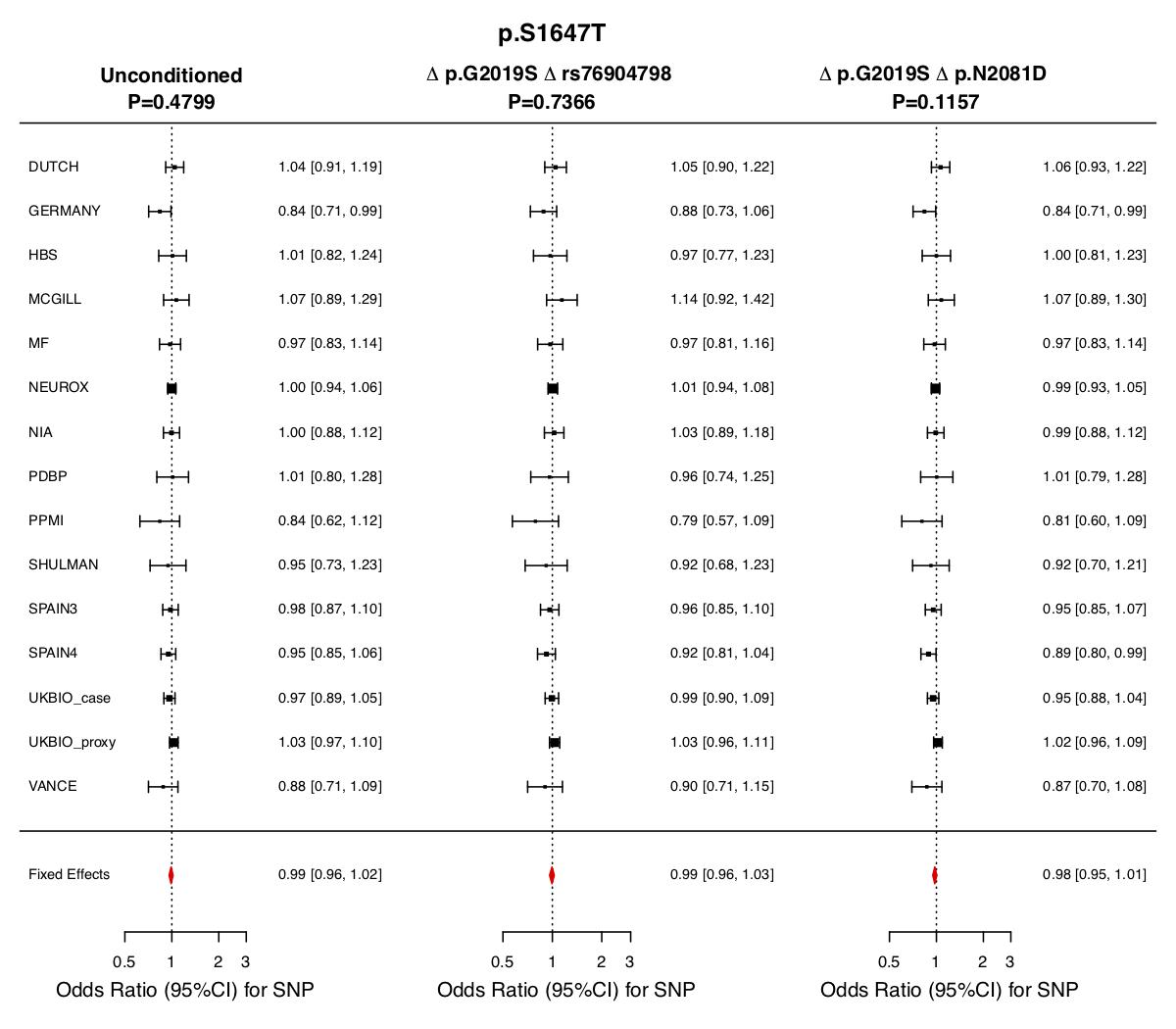

**Supplementary Figure 5.** Meta-analysis of p.S1647T in the included datasets excluding (from left to right) 1) no samples 2) carriers of rs76904798 and p.G2019S and 3) carriers of p.N2081D and p.G2019S.

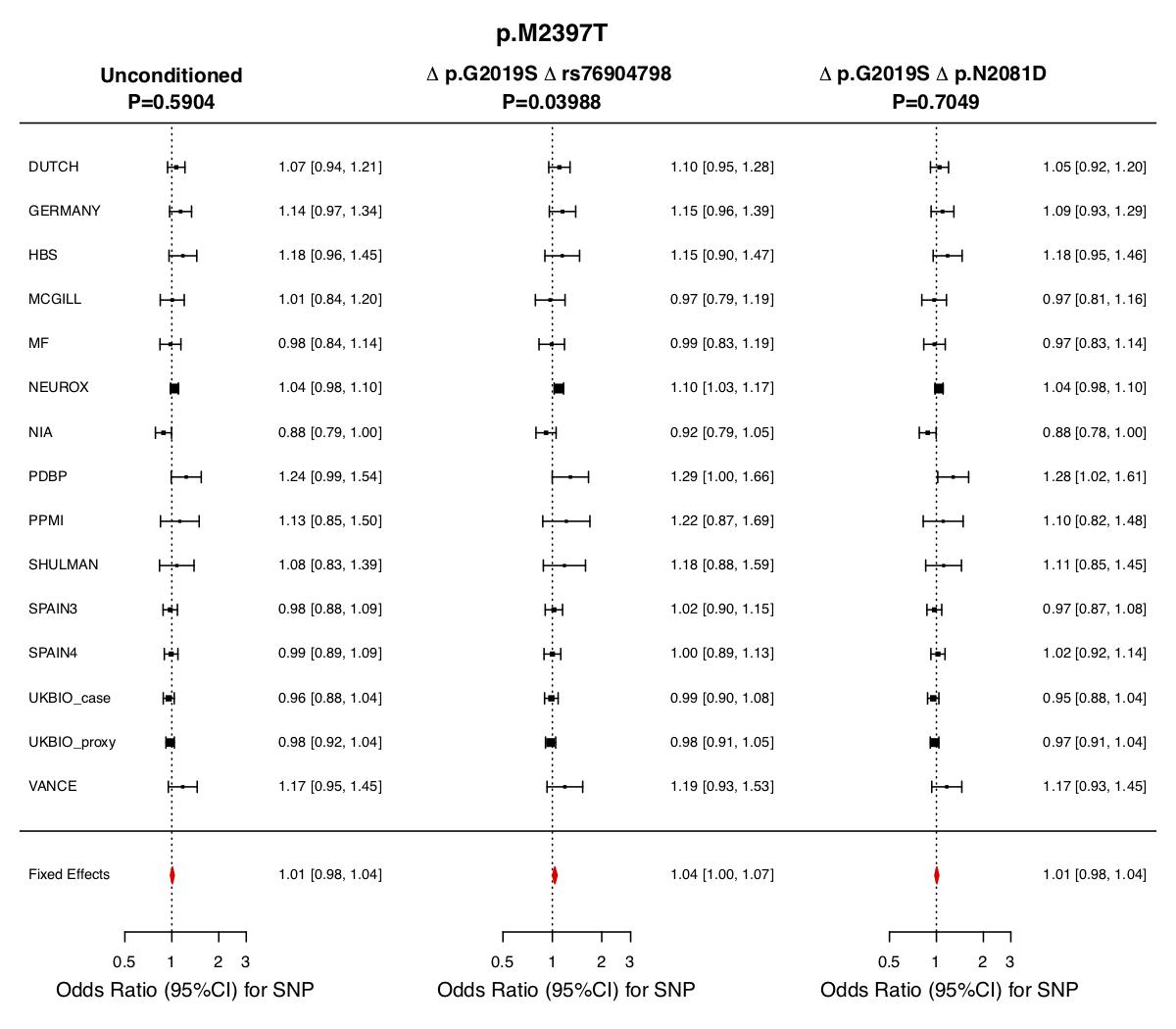
**Supplementary Figure 6.** Meta-analysis of p.M2397T in the included datasets excluding (from left to right) 1) no samples 2) carriers of rs76904798 and p.G2019S and 3) carriers of p.N2081D and p.G2019S.

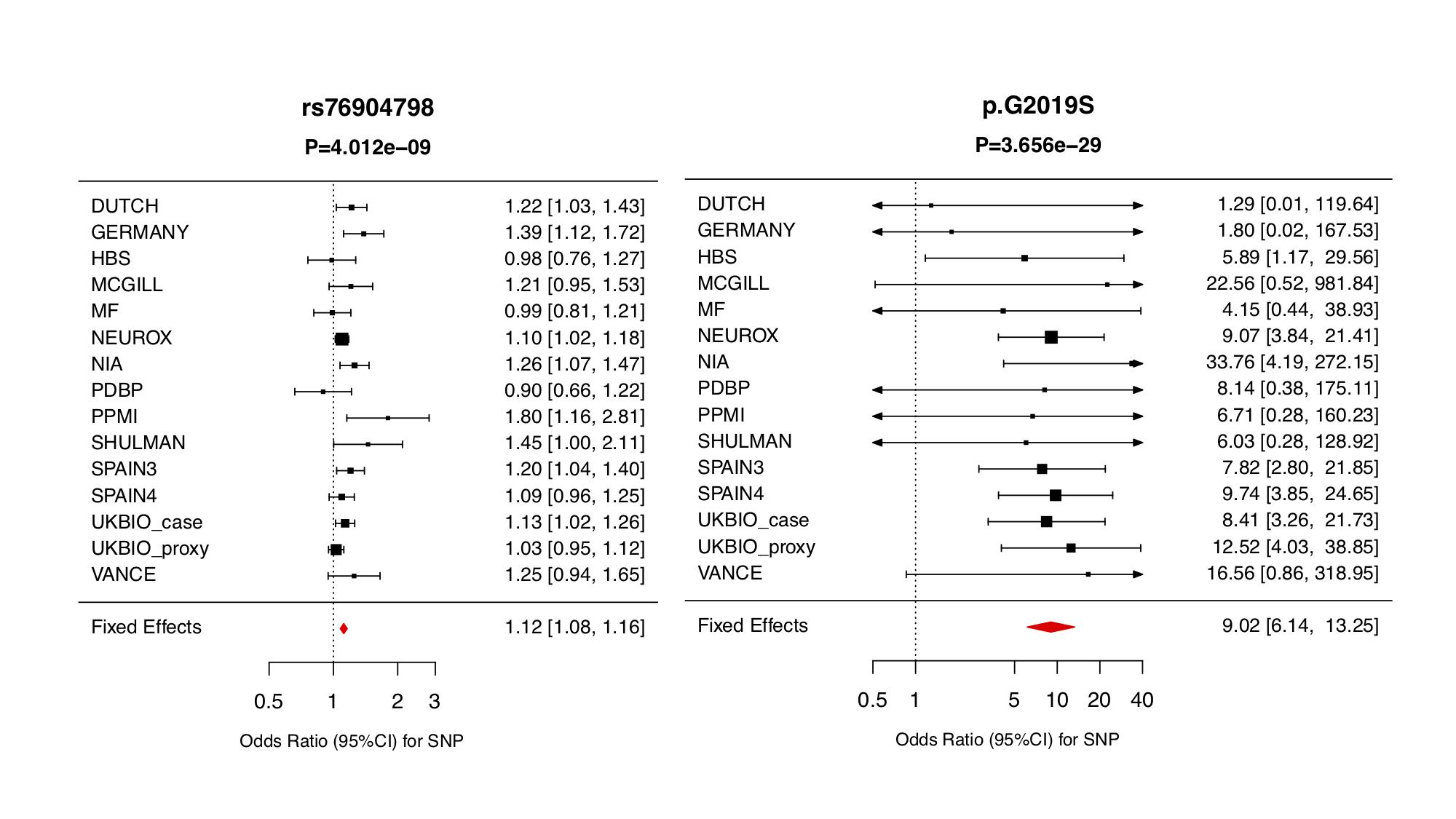

**Supplementary Figure 7.** Meta-analysis of (from left to right) 1) p.G2019S and 2) rs76904798 in the included datasets.

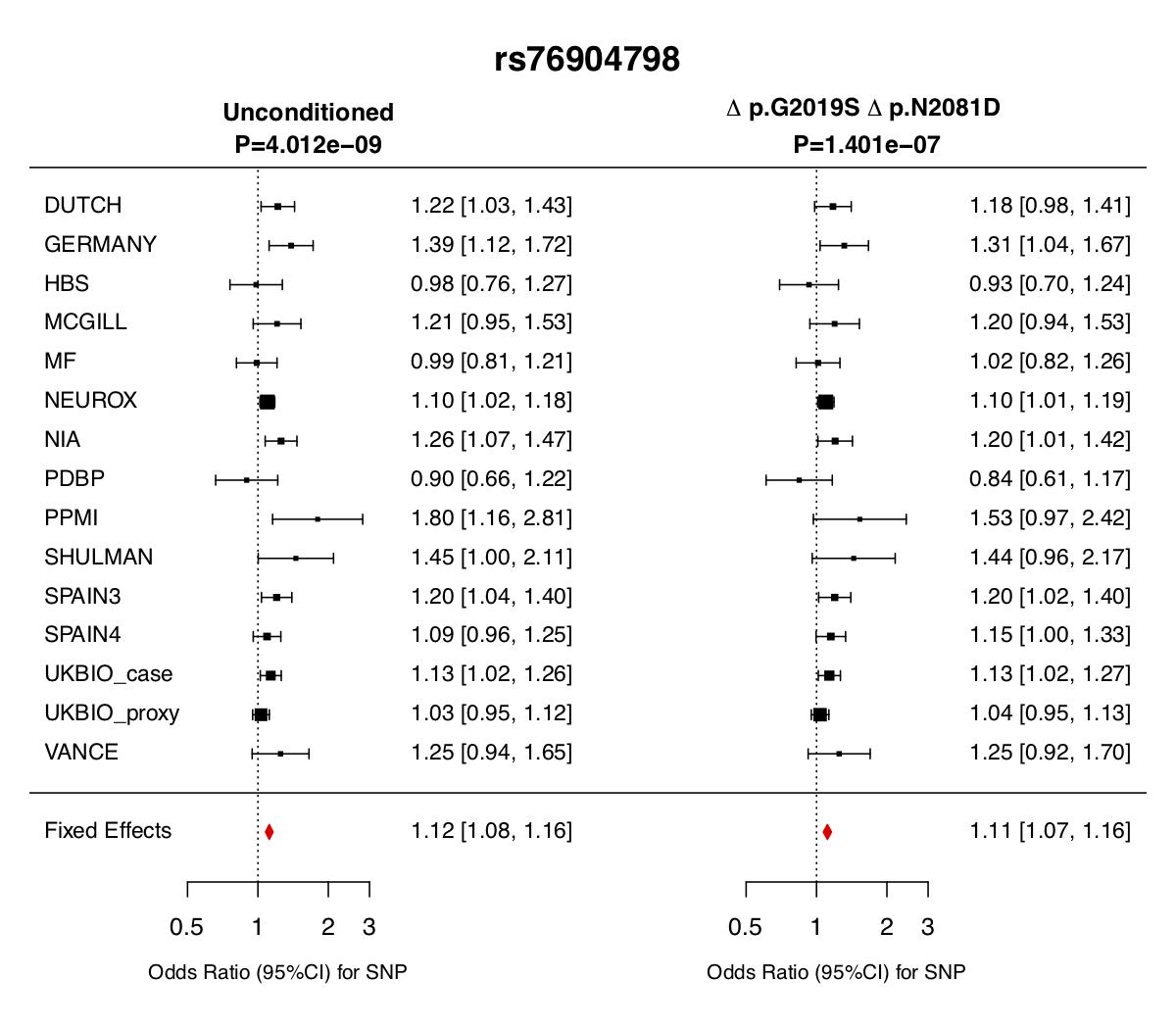

**Supplementary Figure 8.** Meta-analysis of rs76904798 in the included datasets excluding (from left to right) 1) no samples and 2) carriers of p.N2081D and p.G2019S.

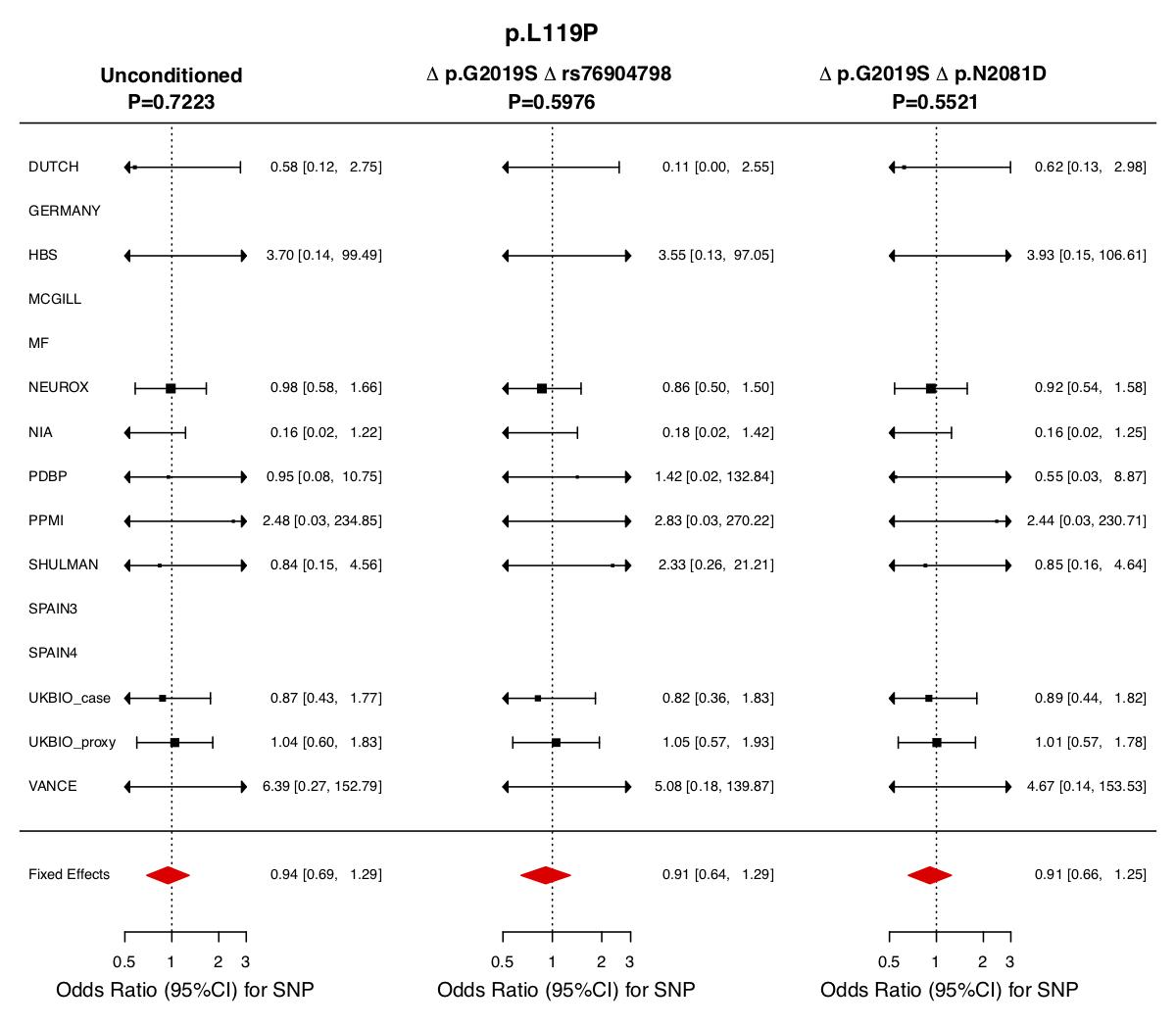

**Supplementary Figure 9.** Meta-analysis of p.L119P in the included datasets excluding (from left to right) 1) no samples 2) carriers of rs76904798 and p.G2019S and 3) carriers of p.N2081D and p.G2019S.

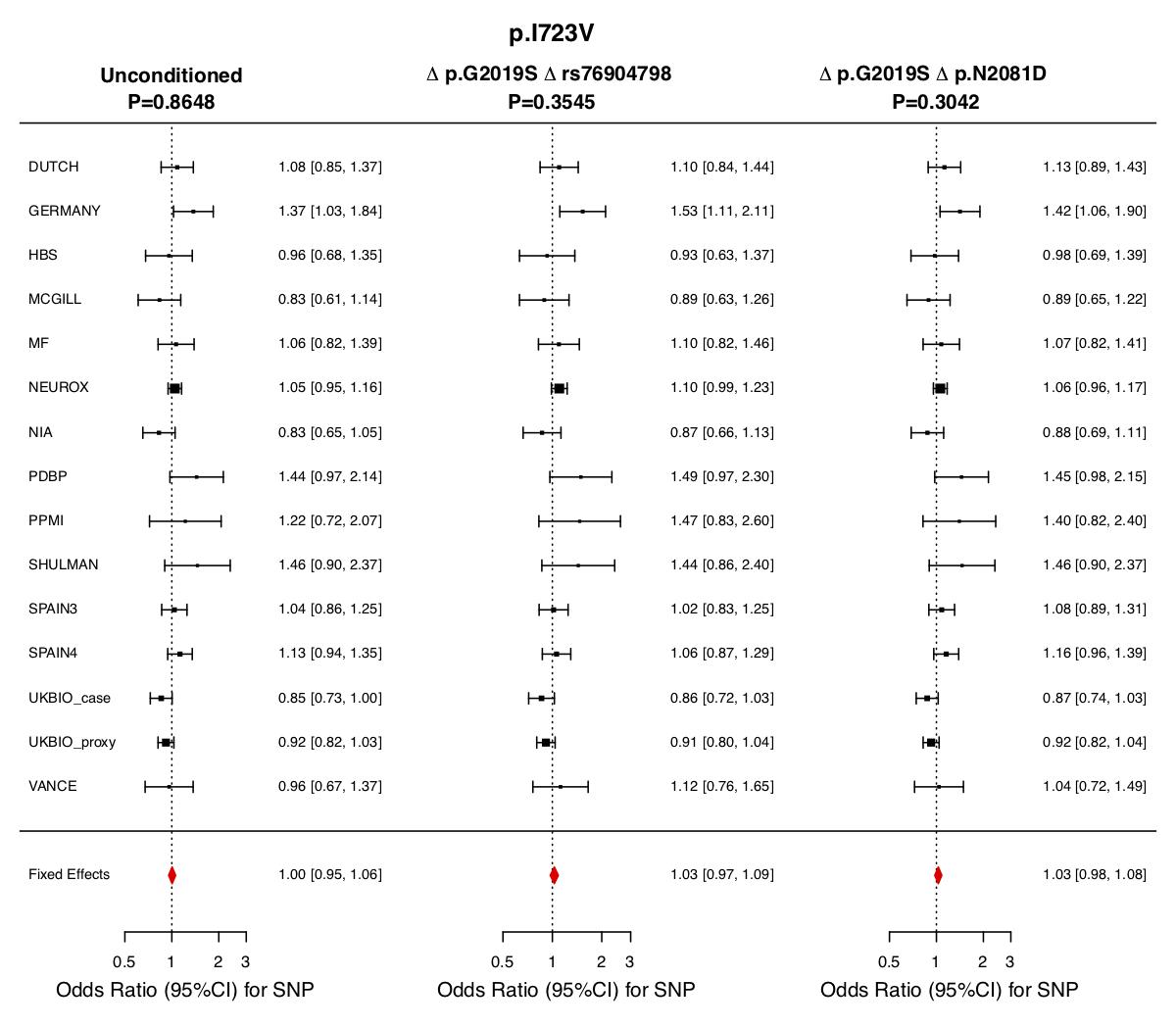

**Supplementary Figure 10.** Meta-analysis of p.I723V in the included datasets excluding (from left to right) 1) no samples 2) carriers of rs76904798 and p.G2019S and 3) carriers of p.N2081D and p.G2019S.

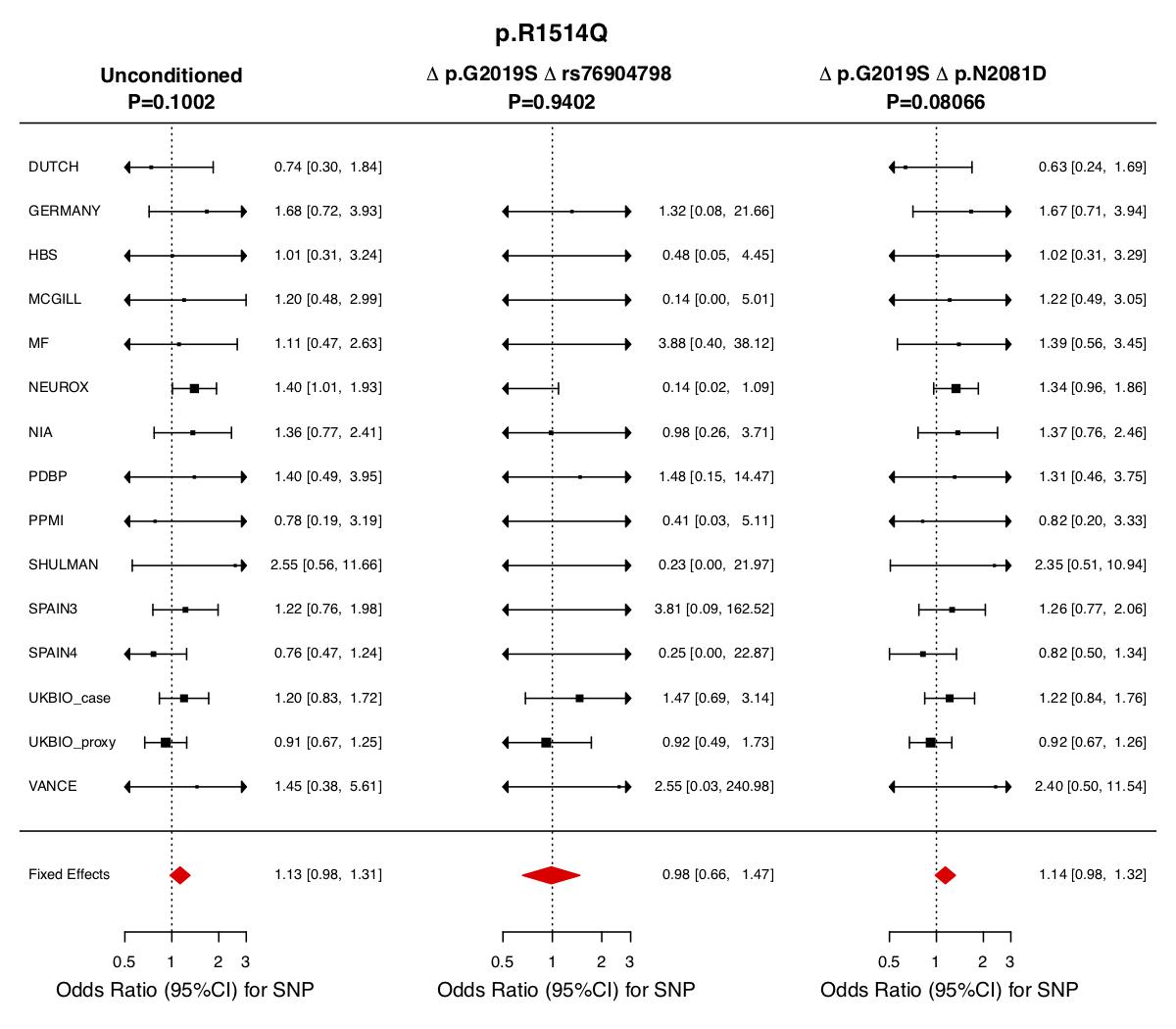

**Supplementary Figure 11.** Meta-analysis of p.R1514Q in the included datasets excluding (from left to right) 1) no samples 2) carriers of rs76904798 and p.G2019S and 3) carriers of p.N2081D and p.G2019S.

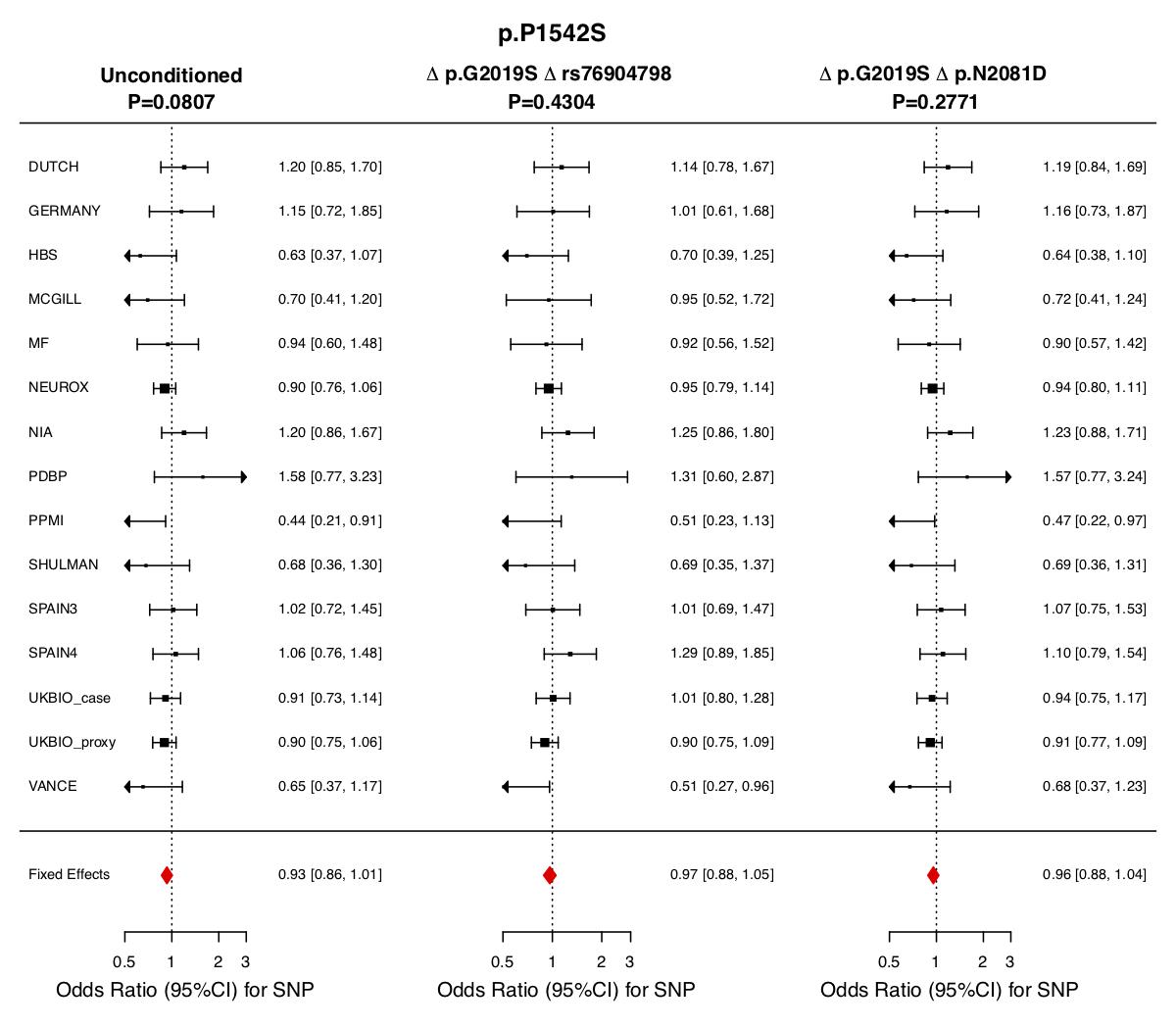
**Supplementary Figure 12.** Meta-analysis of p.P1542S in the included datasets excluding (from left to right) 1) no samples 2) carriers of rs76904798 and p.G2019S and 3) carriers of p.N2081D and p.G2019S.

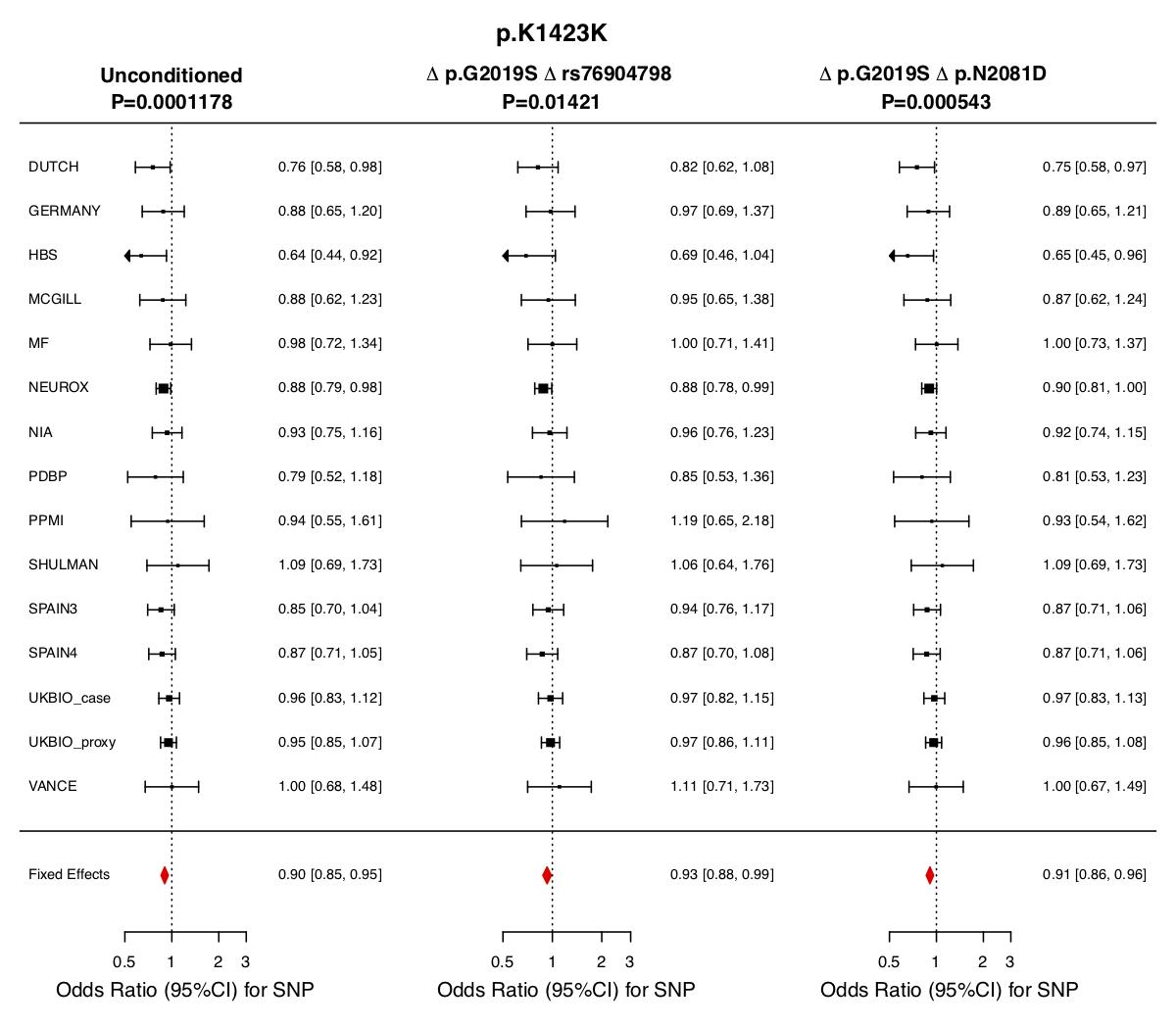

**Supplementary Figure 13.** Meta-analysis of p.K1423K in the included datasets excluding (from left to right) 1) no samples 2) carriers of rs76904798 and p.G2019S and 3) carriers of p.N2081D and p.G2019S.

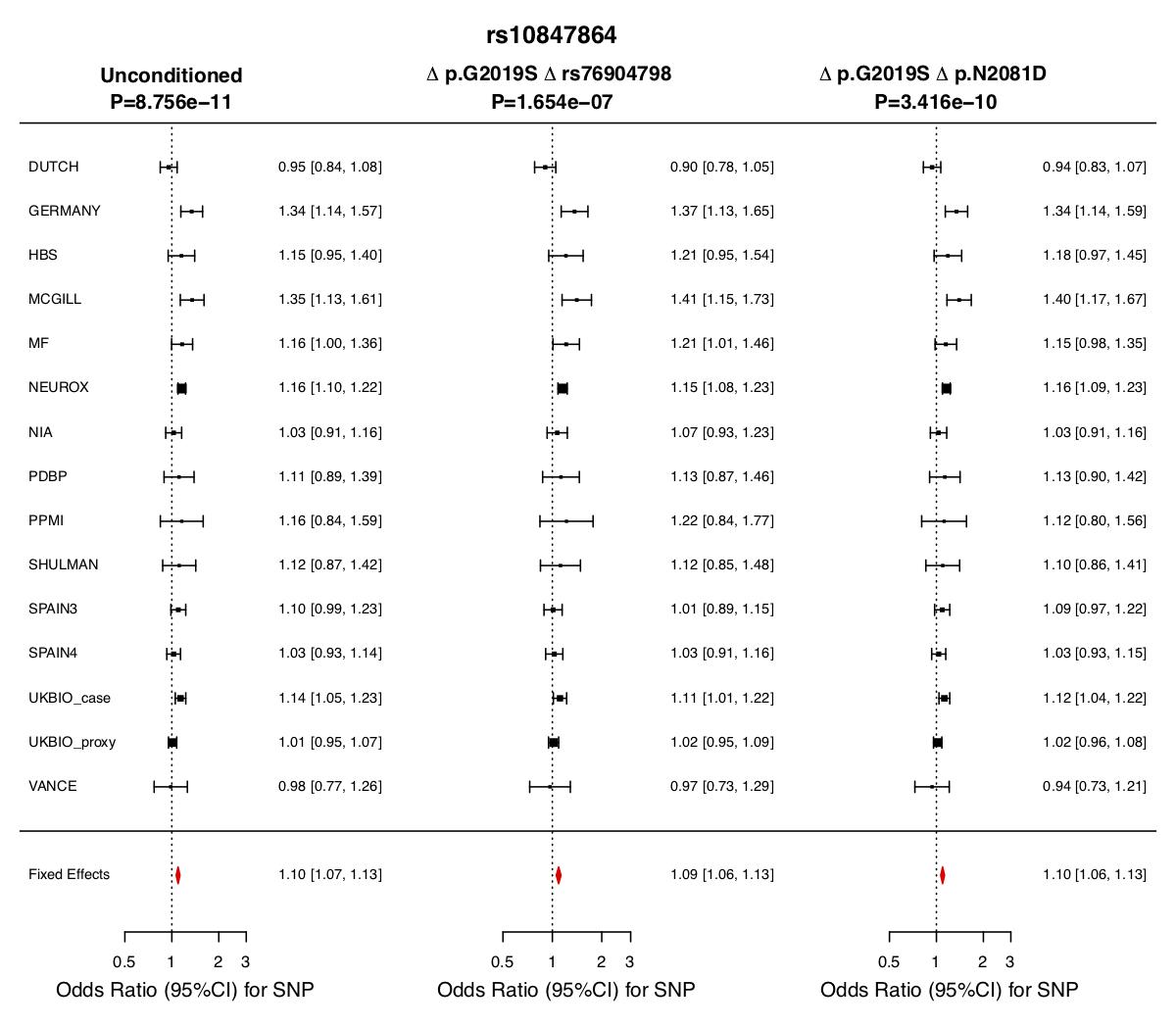

**Supplementary Figure 14.** Meta-analysis of rs10847864 in the included datasets excluding (from left to right) 1) no samples 2) carriers of rs76904798 and p.G2019S and 3) carriers of p.N2081D and p.G2019S.

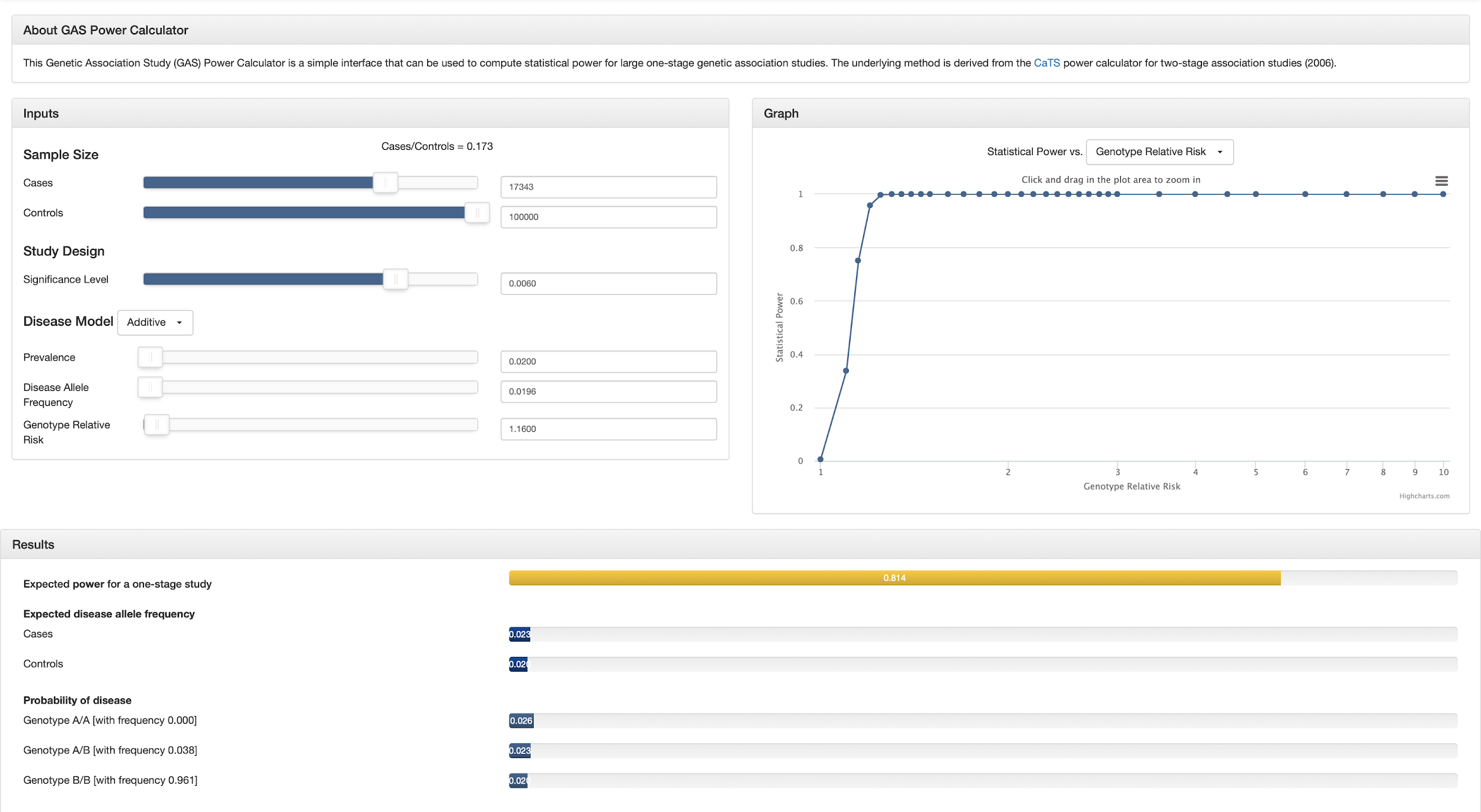
**Supplementary Figure 15.** GAS Power Calculator interface for LRRK2 p.M1646T in the conditional analysis. The left panel shows the input parameters, the right panel shows the genotype relative risks detected at a given power level with these input parameters, and the bottom panel shows the results of the power calculation.

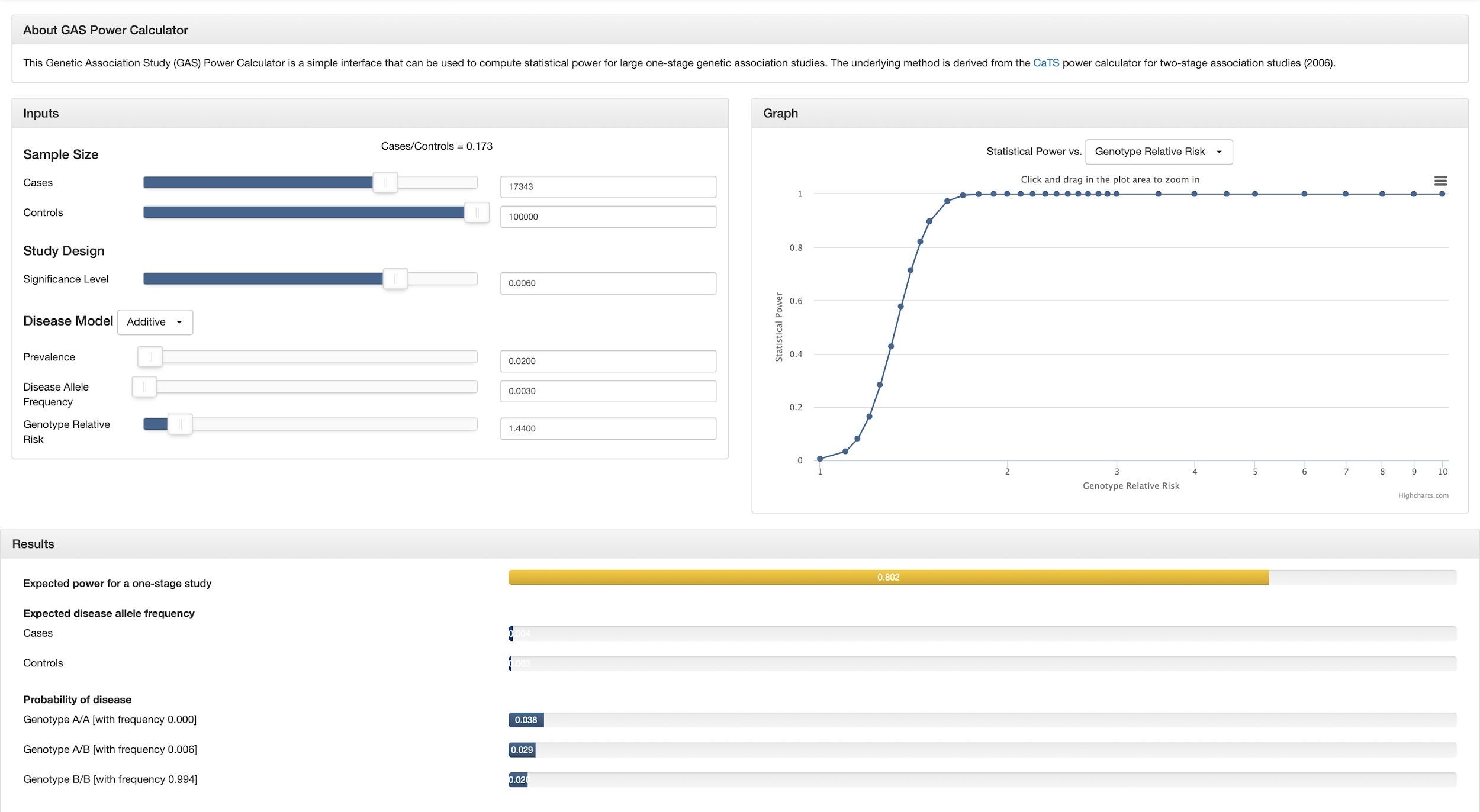
**Supplementary Figure 16.** GAS Power Calculator interface for LRRK2 p.N2081D in the conditional analysis. The left panel shows the input parameters, the right panel shows the genotype relative risks detected at a given power level with these input parameters, and the bottom panel shows the results of the power calculation.

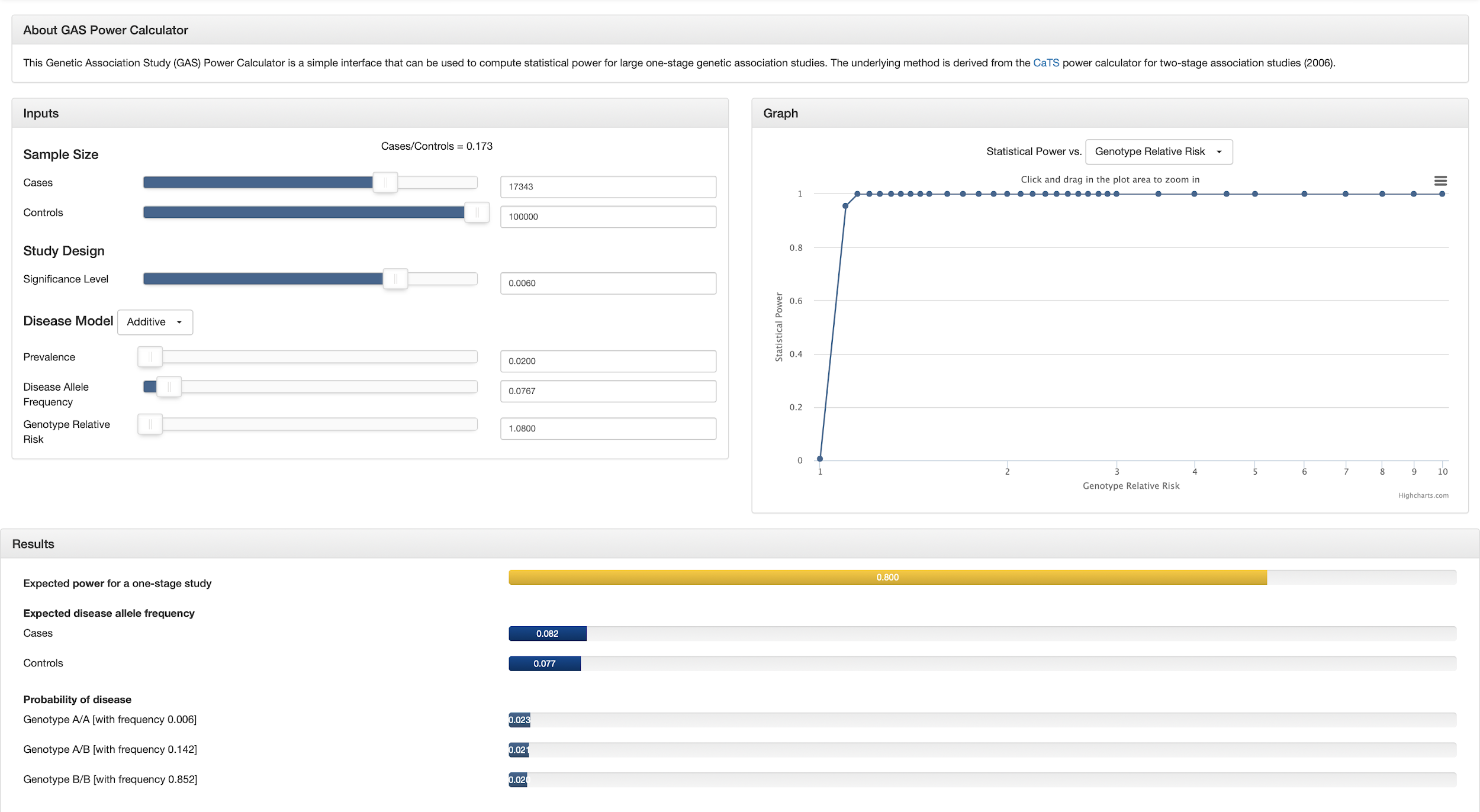
**Supplementary Figure 17.** GAS Power Calculator interface for LRRK2 p.N551K in the conditional analysis. The left panel shows the input parameters, the right panel shows the genotype relative risks detected at a given power level with these input parameters, and the bottom panel shows the results of the power calculation. Since p.N551K has demonstrated a protective association with PD, the resulting relative risk was modified as follows: the natural logarithm was taken, multiplied by -1, and exponentiated.

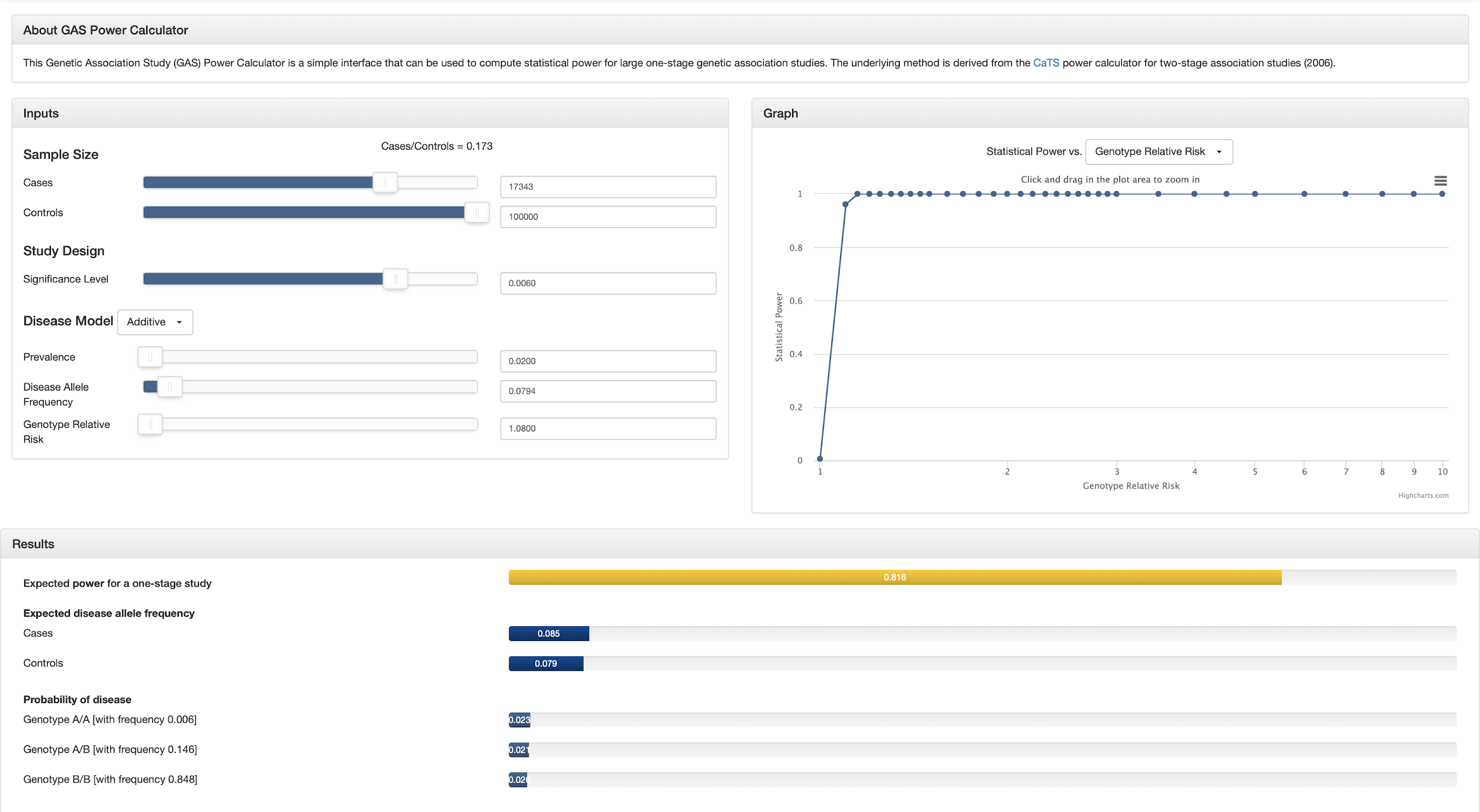
**Supplementary Figure 18.** GAS Power Calculator interface for LRRK2 p.R1398H in the conditional analysis. The left panel shows the input parameters, the right panel shows the genotype relative risks detected at a given power level with these input parameters, and the bottom panel shows the results of the power calculation. Since p.R1398H has demonstrated a protective association with PD, the resulting relative risk was modified as follows: the natural logarithm was taken, multiplied by -1, and exponentiated.

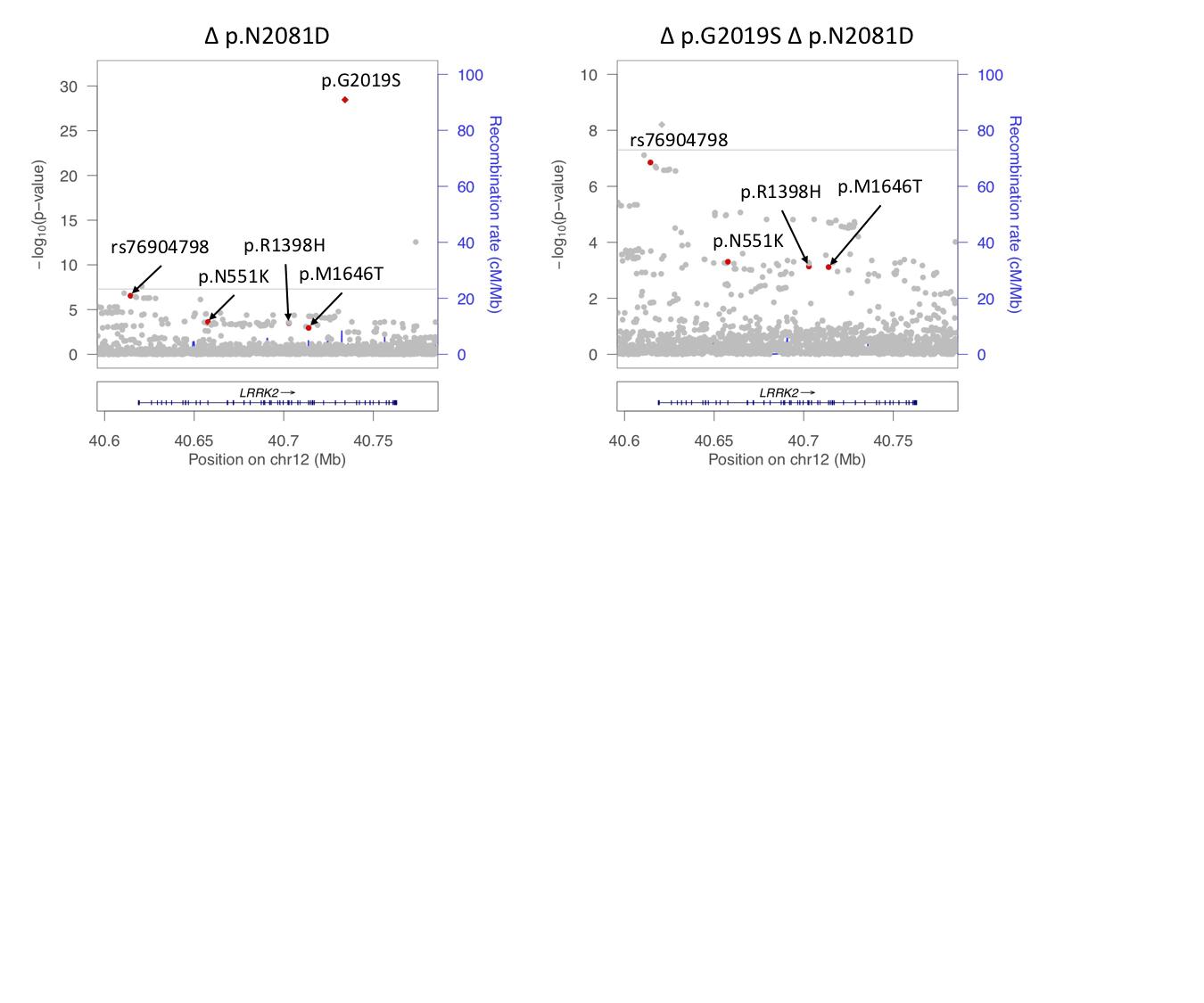
**Supplementary Figure 19.** LocusZoom plot of *LRRK2* association with Parkinson’s disease risk conditioned on p.N2081D. The left panel shows the association signal at the *LRRK2* locus in the IPDGC and UK Biobank meta-analysis conditioned on p.N2081D, and the right panel conditions on both p.G2019S and p.N2081D. The LRRK2 variants p.N551K, p.R1398H, p.M1646T, p.G2019S and rs76904798 are indicated by red dots.
