## Supplementary Methods for "*LRRK2* coding variants and the risk of Parkinson’s disease"

**Quality control and principal component calculation**

*International Parkinson Disease Genomics Consortium genotyping data*

Quality control was done using PLINKv1.9 ^1^. Genotypes were filtered for exclusion using the following criteria: minor allele frequency (MAF) < 1%, genotype call rate < 95%, Hardy‐Weinberg equilibrium P-values of < 1.0E‐4 in controls, and non-random missingness by case/control or haplotype at P-values < 1.0E‐4. Participants were filtered to exclude high missingness (sample call rate < 95%), sex discordance, extreme heterozygosity (F statistics >0.15 or <-0.15) and participants with genetically ascertained relatedness on a level closer than first cousin (pair‐wise kinship coefficients > 0.125). Quality control scripts for filtering can be found at <https://github.com/neurogenetics/GWAS-pipeline>. Principal components (PCs) were calculated from non-imputed genotype data for each dataset using PLINKv1.9 after excluding variants with MAF < 1%, genotype call rate < 85%, and Hardy‐Weinberg equilibrium P-values < 1.0E‐6. PCs were calculated after pruning the remaining variants using a 50-kb window, a 5 SNP shift per window and an r^2^ threshold of 0.5.

*UK Biobank data*

Genotypes were filtered using PLINKv2.0 to exclude variants with a genotype call rate < 90%, and Hardy‐Weinberg equilibrium P-values of < 1.0E‐6 ^2^. QC steps for the exclusion of participants included filtering for European ancestry outliers, relatedness on a level closer than cousin, and sample call rate < 90%. Principal components (PCs) were generated from non-imputed genotype data after filtering using PLINKv1.9 to exclude variants with MAF < 5%, genotype call rate < 99% and Hardy‐Weinberg equilibrium P-values < 5.0E‐6 ^1^. After pruning the remaining variants with a 1000-kb window, a 10 SNP shift per window and an r^2^ threshold of 0.02, PCs were calculated using FlashPCA ^3^.

**Statistical power calculations**

We performed power calculations for the conditional analysis using the GAS Power Calculator ^4^. The number of cases/controls in the IPDGC and UK Biobank datasets after removing carriers of p.G2019S and rs76904798 were 17,343 (12,504 PD cases + 1/2 x 9,679 proxy-cases) and 100,000 (maximum allowed). The significance level used was 0.006, prevalence of PD was 0.02, and disease allele frequency varied depending on the MAF of the variant in the combined IPDGC and UK Biobank datasets used in the conditional analysis (Table 1). The disease prevalence estimate of 2% used for power calculations was somewhat generous but had a very minor impact on the resulting relative risk compared to the more conservative prevalence estimate of 0.5%. Statistical power was calculated using the additive model. Images of the GAS Power Calculator interface are presented in Supplementary Figures 3-6.

### References

1.	Purcell S, Neale B, Todd-Brown K, et al. PLINK: a tool set for whole-genome association and population-based linkage analyses. Am. J. Hum. Genet. 2007;81(3):559–575.

2.	Chang CC, Chow CC, Tellier LC, et al. Second-generation PLINK: rising to the challenge of larger and richer datasets. Gigascience 2015;4:7.

3.	Abraham G, Inouye M. Fast principal component analysis of large-scale genome-wide data. PLoS One 2014;9(4):e93766.

4.	[Johnson JL, Abecasis GR. GAS Power Calculator: web-based power calculator for genetic association studies [Internet]. Cold Spring Harbor Laboratory 2017;164343.[cited 2021 Feb 24 ] Available from:](http://paperpile.com/b/wi6Vit/QadW) <https://www.biorxiv.org/content/10.1101/164343v1.abstract>
